# Efficacy of Proxalutamide in Hospitalized COVID-19 Patients: A Randomized, Double-Blind, Placebo-Controlled, Parallel-Design Clinical Trial

**DOI:** 10.1101/2021.06.22.21259318

**Authors:** Flávio Adsuara Cadegiani, Daniel do Nascimento Fonseca, John McCoy, Ricardo Ariel Zimerman, Fatima Nadeem Mirza, Michael do Nascimento Correia, Renan Nascimento Barros, Dirce Costa Onety, Karla Cristina Petruccelli Israel, Brenda Gomes de Almeida, Emilyn Oliveira Guerreiro, José Erique Miranda Medeiros, Raquel Neves Nicolau, Luiza Fernanda Mendonça Nicolau, Rafael Xavier Cunha, Maria Fernanda Rodrigues Barroco, Patrícia Souza da Silva, Gabriel de Souza Ferreira, Flavio Renan Paula da Costa Alcântara, Ângelo Macedo Ribeiro, Felipe Oliveira de Almeida, Adailson Antonio de Souza Silva, Suzyane Serfaty do Rosario, Raysa Wanzeller de Souza Paulain, Alessandra Reis, Marissa Li, Claudia Elizabeth Thompson, Gerard J. Nau, Carlos Gustavo Wambier, Andy Goren

## Abstract

**Background:** Severe acute respiratory syndrome coronavirus 2 (SARS-CoV-2) infectivity is mediated by the androgen-promoted protease, transmembrane protease, serine 2 (TMPRSS2). Previously, we have shown that treatment with proxalutamide, a non-steroidal androgen receptor antagonist, accelerates viral clearance and clinical remission in outpatients with coronavirus disease 2019 (COVID-19) compared to placebo. The effects in hospitalized COVID-19 patients were unknown.

**Methods:** Men and women hospitalized but not requiring mechanical ventilation were randomized (1:1 ratio) to receive 300 mg of proxalutamide per day or placebo for 14 days. The study was conducted at eight sites in the state of Amazonas, Brazil. The primary outcome measure was the clinical status (8-point ordinal scale) at 14-days post-randomization. The primary efficacy endpoint was the 14-day recovery ratio (alive hospital discharge [scores 1, 2]).

**Findings:** A total of 645 patients were randomized (317 received proxalutamide, 328 placebo) and underwent intention-to-treat analysis. The 14-day median ordinal scale score in the proxalutamide group was 1 (interquartile range [IQR]=1–2) versus 7 (IQR=2–8) for placebo, P<0.001. The 14-day recovery rate was 81.4% for proxalutamide and 35.7% for placebo (recovery ratio, 2.28; 95% CI 1.95–2.66 [P<0.001]). The 28-day all-cause mortality rate was 11.0% for proxalutamide versus 49.4% for placebo (hazard ratio, 0.16; 95% CI 0.11–0.24). The median post-randomization time to recovery was 5 days (IQR=3– 8) for proxalutamide versus 10 days (IQR=6–15) for placebo.

**Interpretation:** Hospitalized COVID-19 patients not requiring mechanical ventilation receiving proxalutamide had a 128% higher recovery rate than those treated with placebo. All-cause mortality was reduced by 77.7% over 28 days. (ClinicalTrials.gov number, NCT04728802).

## Introduction

Severe acute respiratory syndrome coronavirus 2 (SARS-CoV-2) and the resulting coronavirus disease 2019 (COVID-19) has claimed 2.6 million lives globally since it first emerged in December of 2019.^1^ SARS-CoV-2 infects type II pneumocytes in the human lung and endothelial cells by anchoring to angiotensin-converting enzyme 2 (ACE2) receptors. Prior to binding to ACE2, spike proteins on the viral surface undergo structural modification via endogenous transmembrane protease, serine 2 (TMPRSS2).^2^ Because of this modification, Hoffman, et al. proposed that inhibitors of TMPRSS2 would limit SARS-CoV-2 infection.^2^ The TMPRSS2 promoter includes a 15 base pair androgen response element.^3^ This led our group to hypothesize that antiandrogen drugs would be good candidates for limiting SARS-CoV-2 entry into cells.^4^ While the mechanism of action of antiandrogens against SARS-CoV-2 is not fully elucidated, pre-clinical studies have shown that nonsteroidal antiandrogens down regulate TMPRSS2^5^ and inhibit viral replication in human cell culture.^6,7^

Proxalutamide is a second-generation nonsteroidal androgen receptor antagonist that is more potent than other antiandrogen compounds such as enzalutamide or bicalutamide.^8^ These compounds competitively inhibit androgen binding, block androgen receptor nuclear translocation, and prevent their binding to DNA.^9^ We previously studied the use of proxalutamide, in SARS-CoV-2 positive men in an outpatient setting. In a double-blinded, placebo-controlled, randomized clinical trial, men treated with proxalutamide (200 mg/day) demonstrated significantly reduced hospitalization rates, accelerated improvements of COVID-19 symptoms, and accelerated viral clearance.^10^ Proxalutamide also reduced the duration of COVID-19 in both men and women diagnosed with COVID-19 in the outpatient setting.^11^ Here, we evaluated the efficacy of proxalutamide compared to the usual care in hospitalized men and women with COVID-19.

## Methods

### Trial Design, Setting and Locations

This was a double-blinded, randomized, placebo-controlled, prospective, two-arm trial. The trial was conducted at eight centers in six cities of the state of Amazonas, Brazil from February 1 to April 15, 2021, including enrollment and follow-up.

### Eligibility criteria

Inclusion criteria: Men and women hospitalized due to COVID-19 with a previously confirmed positive test for SARS-CoV-2 within 7 days prior to randomization. SARS-CoV-2 status was determined by real-time reverse transcription polymerase chain reaction (rtPCR) testing following the Cobas SARS-CoV-2 rtPCR kit test protocol (Roche, USA).

Exclusion criteria included mechanical ventilation at the time of randomization, a history of congestive heart failure class III or IV (New York Heart Association), immunosuppression, alanine transferase (ALT) above five times ULN (> 250 U/L), creatinine above 2.5 mg/ml, or a calculated eGFR was below 30 ml/min. Patients currently using any antiandrogen medications were also excluded. In female patients, those that were pregnant, breastfeeding, or were planning to become pregnant within 90 days were also excluded.

Patients were randomized to receive either proxalutamide 300 mg/day plus usual care or a placebo plus usual care for 14 days in a 1:1 ratio. If patients were discharged before 14 days, they were instructed to continue treatment. Therapy compliance was monitored daily.

### Procedures

The COVID-19 8-point ordinal scale was used for screening (day 0) as well as daily clinical assessments of patients on days 1-14 (or inpatient after day 14), as well as day 21, and day 28 (if outpatient), resulting in a maximum of 17 data points for each patient. The clinical score is defined as: 8. Death; 7. Hospitalized, on invasive mechanical ventilation; 6. Hospitalized, on non-invasive ventilation or high flow oxygen devices; 5. Hospitalized, requiring supplemental oxygen; 4. Hospitalized, not requiring supplemental oxygen-requiring ongoing medical care (COVID-19 related or otherwise); 3. Hospitalized, not requiring supplemental oxygen - no longer requires ongoing medical care; 2. Not hospitalized, limitation on activities; and 1. Not hospitalized, no limitations on activities.^12^

Participants discharged from the hospital were evaluated by investigators in follow-up appointments whenever possible, or by daily phone calls. Upon hospital discharge, patients were instructed to contact the same hospital and the local study team in case of relapse or new symptoms. Hospital readmissions were actively surveilled in all sites.

Baseline characteristics, previous medical history, and concomitant medications were recorded for each patient. Proxalutamide 300 mg/day or placebo plus usual care was given for 14 days, even when COVID-19 remission occurred before this period. Usual care included medications such as enoxaparin, colchicine, methylprednisolone, dexamethasone, or antibiotic therapy if necessary.

Before the onset of the trial, a random sequence was created using a web-based randomization software^13^ using 4, 6 and 8 block sizes and a list length for 662 treatments. The randomization sequence and the allocation concealment were performed remotely from the patient recruiting centers and it was not stratified by site. Pre-packing of tablets of either active or placebo group was manufactured to appear identical (Kintor Pharmaceuticals Ltd. Suzhou, China). The procedure is detailed in the **Supplementary Appendix**.

The local investigators who were directly involved with patient care, other healthcare providers, and patients were kept blinded to the group assignments until all patients completed the 28-day post-randomization period and the data was locked. The principal investigator was unblinded for data and safety analysis in March 2021. The interim analysis was conducted on March 10^th^ 2021, when the primary outcome measure was reported for a total of 590 patients.

### Outcomes and Statistical Analysis

The primary outcome measure was the 8-point COVD-19 ordinal scale at post-randomization day 14. The primary efficacy endpoint measure was the overall recovery ratio, which was calculated from recovery rates in each group. Recovery was defined as achieving alive hospital discharge (scores 1 and 2). The secondary outcome measures included recovery rate and all-cause mortality rate (score 8) and respective risk ratios at post-randomization day 28; all-cause mortality hazard ratio; median hospitalization time; and median post-randomization time to recover (alive hospital discharge). Subgroup analysis included sex and baseline scores.

An intention-to-treat protocol was used for data analysis. The Wilcoxon Rank Sum test was used to assess the differences of the ordinal scale scores at 14 and 28 days. Risk ratios were calculated to measure the effects of proxalutamide versus placebo on the recovery and all-cause mortality rates. Additional analysis included the recovery and mortality risk ratios by gender, baseline COVID-19 ordinal score, and hospital site. To evaluate the all-cause mortality and recovery over the 28-days post-randomization observation period, we used Kaplan-Meier’s survivor function for estimates of proportion surviving and failure function for estimates of alive hospital discharges. Cox proportional hazards model was used to calculate hazard ratio (HR) for all-cause mortality over 28 days and its 95% confidence interval (CI). Graphical assessment and Kaplan-Meier versus predicted survival showed that of the proportional-hazards assumption has not been violated, **Figure S3**.

The sample size was calculated to be able to detect a difference of approximately 14% in the overall recovery rate at 14 days (risk ratio of 1.36) with a power of 90% and a type I error of 5%, over an estimated 39% recovery rate for the placebo group (based on the protocol for NCT04280705 [scenario 4]), and 3.5% non-compliance/cross over for each group. Statistical significance was set at P<0.05 and no correction for multiple comparisons was performed. Stata/SE version 16.1 for Mac (StataCorp LLC, College Station, TX, USA) was used to perform all statistical analysis.

## Results

### Patients

Between February 1 and March 17, 2021, 697 patients were assessed for eligibility, including 396 males (56.8%) and 301 females (43.2%). At screening, twelve patients did not meet the eligibility criteria as they had kidney failure or liver abnormal enzymes (1.7%) and forty patients declined enrollment (5.7%). Of the 697 patients initially assessed, 645 underwent randomization. Three hundred seventeen patients were randomized to receive proxalutamide in addition to usual care, including 184 males (58.0%) and 133 females (42.0%). Three hundred twenty-eight were assigned to receive placebo in addition to usual care, including 182 males (55.5%) and 146 females (44.5%). After randomization, 29 of the 317 (9.1%) patients receiving proxalutamide and 36 of the 328 (10.9%) receiving placebo did not complete the trial protocol, the reasons are described in **Figure 1**.

**Figure 1.**
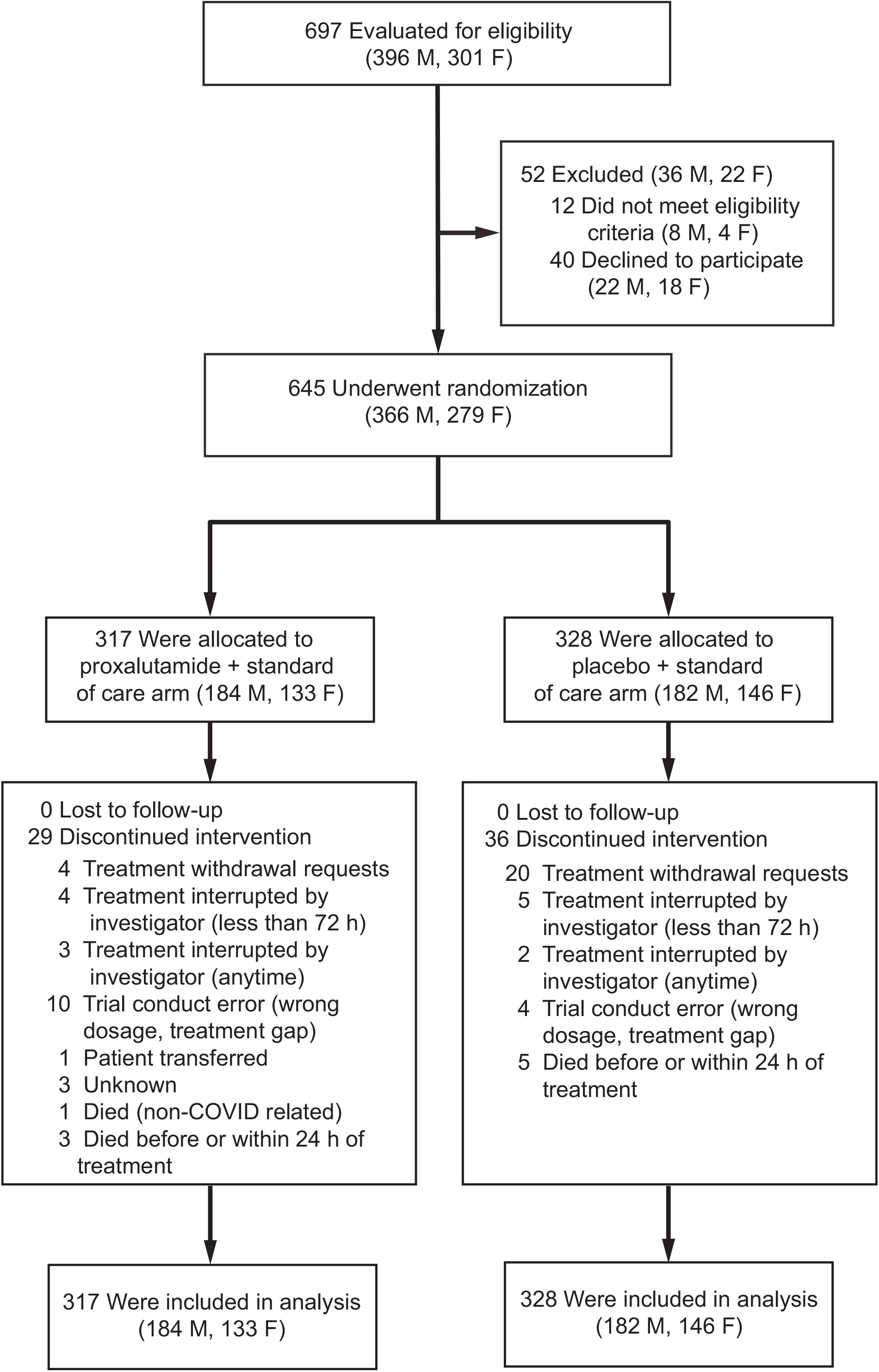
Enrollment and randomization of the studied population.

The baseline characteristics of the study populations are described in **Table 1**. The median age was 50 years (interquartile range [IQR], 41 to 62) and 49 years (IQR, 38 to 61) for the proxalutamide and placebo groups, respectively. Patients with a body mass index (BMI) above 30kg/m2, hypertension, type 2 diabetes mellitus (T2DM), and chronic obstructive pulmonary disorder (COPD) were equally distributed between study arms. No comorbidities, one comorbidity, and two or more comorbidities were present in 69.4%, 17.3%, and 13.3% of participants, respectively, and were equally distributed between study arms. Median time from hospitalization to randomization was 2 days (IQR 1 to 4.2) for placebo and 2 days (IQR 1 to 4) for proxalutamide.

**Table 1.** Baseline clinical characteristics, comorbidities, and concomitant medications of the studied population (IQR = interquartile range).

| Characteristic | Overall<br>N=645 | Proxalutamide<br>N=317 | Placebo<br>N=328 |
| --- | --- | --- | --- |
| **Age** |  |  |  |
| **Median – years (IQR)** | 50 (39-61.5) | 50 (41-62) | 49 (38-61) |
| **> 55 yr – no. (%)** | 238 (36.9) | 115 (36.3) | 123 (37.5) |
| **Sex – no. (%)** |  |  |  |
| **Female** | 279 (43.3) | 133 (42.0) | 146 (44.5) |
| **Male** | 366 (56.7) | 184 (58.0) | 182 (55.5) |
| **Body mass index over 30 kg/m<sup>2</sup> – no. (%)** | 53 (8.2) | 28 (8.8) | 25 (7.6) |
| **Hypertension – no. (%)** | 175 (27.1) | 93 (29.3) | 82 (25.0) |
| **Type 2 diabetes mellitus – no. (%)** | 79 (12.2) | 41 (12.9) | 38 (11.6) |
| **Chronic obstructive pulmonary disorder – no. (%)** | 16 (2.5) | 8 (2.5) | 8 (2.4) |
| **Chronic kidney disease – no. (%)** | 0 (0.0) | 0 (0.0) | 0 (0.0) |
| **Coexisting conditions – no. (%)** |  |  |  |
| 0 | 431 (66.8) | 204 (64.4) | 227 (69.2) |
| 1 | 124 (19.2) | 68 (21.4) | 56 (17.1) |
| 2+ | 90 (14.0) | 45 (14.2) | 45 (13.7) |
| **Median time from hospitalization to randomization (IQR) – days** | 2.0 (1.0-4.0) | 2.0 (1.0-4.0) | 2.0 (1.0-4.2) |
| **Score on the Coronavirus Disease 2019 ordinal scale– no. (%)** |  |  |  |
| **3. Hospitalized, not requiring supplemental oxygen - no longer requires ongoing medical care** | 2 (0.3) | 0 (0.0) | 2 (0.6) |
| **4. Hospitalized, not requiring supplemental oxygen, requiring ongoing medical care (COVID-19 related or otherwise)** | 18 (2.8) | 7 (2.2) | 11 (3.3) |
| **5. Hospitalized, requiring supplemental oxygen** | 196 (30.4) | 93 (29.3) | 103 (31.4) |
| **6. Hospitalized, receiving non-invasive ventilation or high flow oxygen devices** | 429 (66.5) | 217 (68.5) | 212 (64.6) |
| **Concomitant medications – no. (%)** |  |  |  |
| Ceftriaxone | 638 (98.9) | 313 (98.7) | 325 (99.1) |
| Colchicine | 407 (63.1) | 201 (63.4) | 306 (93.3) |
| Macrolides (azithromycin, clarithromycin) | 631 (97.8) | 307 (96.8) | 324 (98.8) |
| Glucocorticosteroids (dexamethasone, methylprednisolone) | 645 (100) | 317 (100) | 328 (100) |
| Enoxaparin | 645 (100) | 317 (100) | 328 (100) |
| Omeprazole | 645 (100) | 317 (100) | 328 (100) |

The ordinal scale scores at baseline were 6, 5, and 4 in 66.5%, 30.4%, and 2.8% of the population, respectively. The distribution of ordinal scale score was similar between proxalutamide and placebo arms. Except for colchicine, all concomitant medications were used at similar proportions between the groups. Remdesivir was not a treatment option (emergency use authorization dated March 12, 2021 in Brazil).^14^ Compliance with the 14-day treatment was 91% and 89% for the proxalutamide and placebo arms, respectively. Of note, since the study was conducted in Brazil, the population is ethnically mixed and the majority of patients did not identify themselves as Caucasian, black, or Asian.

### Primary Outcome

At the 14-day timepoint, a lower score distribution was observed for the proxalutamide group than in the placebo group, **Figure 2A**. The median score in the proxalutamide group was 1 (IQR 1 to 2) versus 7 (IQR 2 to 8) for placebo (P<0.001), **Table 2**. The overall 14-day recovery rate for placebo of 35.7% (95% CI 30.7 to 40.1) was lower than for proxalutamide (81.4%; 95% CI 76.7 to 85.3), *P*<0.001. The 14-day recovery ratio was 2.28 (95% CI 1.95 to 2.66 [P<0.001]), which indicates patients who took proxalutamide had a 128% higher recovery rate than those treated with placebo (95% CI 95 to 166%). No interaction effect of the primary outcome and gender was observed, **Table 2** and **Figure 2B**.

**Table 2.** Outcomes according to treatment group in the intention-to-treat analysis. Recovery rates (reaching scores 1 or 2) over 14 and 28 days after randomization. All-cause mortality rate over 28 days. (IQR = interquartile range, CI = confidence interval, HR = Hazard Ratio). Subgroup analysis per sex and per baseline score are presented with 95% CI which have not been adjusted for multiplicity.

| Characteristic | Overall<br>N=645 | Proxalutamide<br>N=317 | Placebo<br>N=328 | *Risk ratio (95% CI)* |
| --- | --- | --- | --- | --- |
| **Day 14 Scores – median (IQR)** | 2 (1-7) | 1(1-2) | 7 (2-8) | P<0.001 |
| **Recovery rate over 14 days, no. (%)** | 375 (58.1) | 258 (81.4) | 117 (35.7) | 2.28 (1.95-2.66) P<0.001 |
| Females | 160 (57.3) | 109 (82.0) | 51 (34.9) | 2.35 (1.85-2.97) |
| Males | 215 (58.7) | 149 (81.0) | 66 (36.3) | 2.23 (1.82-2.74) |
| Baseline Score 3 | 2 (100.0) | - | 2 (100.0) | - |
| Baseline Score 4 | 16 (88.9) | 7 (100.0) | 9 (81.8) | 1.22 (0.93–1.61) |
| Baseline Score 5 | 133 (67.9) | 82 (88.2) | 51 (49.5) | 1.78 (1.45–2.19) |
| Baseline Score 6 | 224 (52.2) | 169 (77.9) | 55 (25.9) | 3.00 (2.37–3.81) |
| **Recovery rate over 28 days no. (%)** | 426 (66.0) | 271 (85.5) | 155 (47.3) | 1.81 (1.60-2.05) |
| Females | 188 (67.4) | 117 (88.0) | 71 (48.6) | 1.81 (1.51-2.16) |
| Males | 238 (65.0) | 154 (83.7) | 84 (46.2) | 1.81 (1.53-2.15) |
| Baseline Score 4 | 16 (88.9) | 7 (100.0) | 9 (81.8) | 1.22 (0.93–1.61) |
| Baseline Score 5 | 152 (77.6) | 86 (92.5) | 66 (64.1) | 1.44 (1.23–1.69) |
| Baseline Score 6 | 256 (59.7) | 178 (82.0) | 78 (36.8) | 2.23 (1.85–2.69) |
| **All-cause mortality rate over 28 days, no. (%)** | 197 (30.5) | 35 (11.0) | 162 (49.4) | 0.22 (0.16-0.31)<br>HR 0.16 (0.11-0.24)) |
| Females | 83 (29.7) | 12 (9.0) | 71 (48.6) | 0.18 (0.11-0.33) |
| Males | 114 (31.1) | 23 (12.5) | 91 (50.0) | 0.25 (0.17-0.38) |
| Baseline Score 4 | 2 (11.1) | 0 (0.0) | 2 (18.2) | 0 |
| Baseline Score 5 | 42 (21.4) | 5 (5.4) | 37 (35.9) | 0.15 (0.06–0.36) |
| Baseline Score 6 | 153 (35.7) | 30 (13.8) | 123 (58.0) | 0.24 (0.17–0.34) |
| **Median hospitalization days (IQR)** | 10 (6-16) | 8 (6-13) | 12 (8-18) |  |
| **Post-randomization time to recover / alive hospital discharge, median days (IQR)** | 6 (4-11) | 5 (3-8) | 10 (6-15) |  |

**Figure 2.**
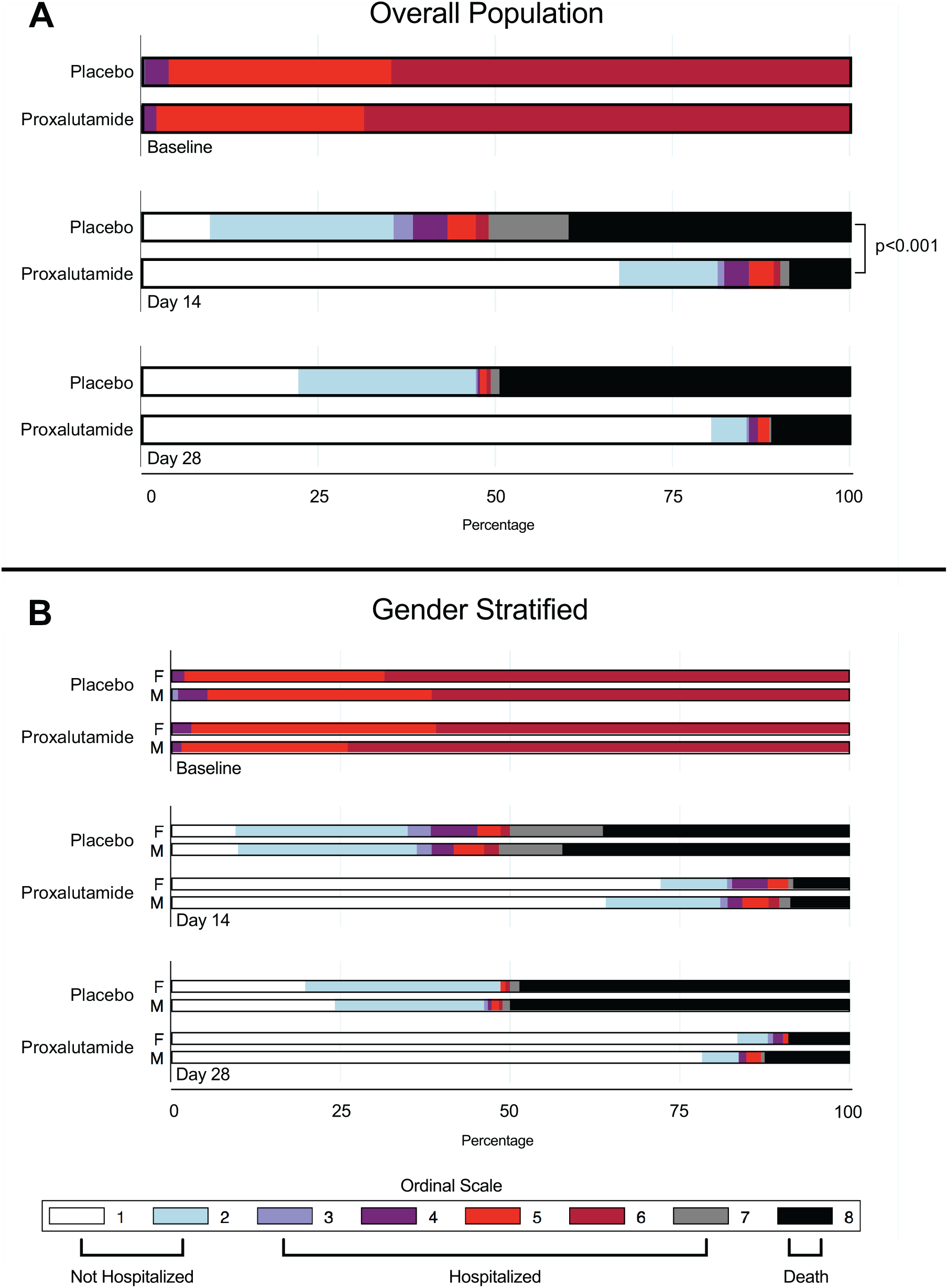
Distribution of the COVID-19 8-Point Ordinal Scale in the Intention-to-Treat population at randomization, day 14, and day 28. Data are presented in the overall population (A) and stratified by gender (B).

### Secondary Outcomes

At the 28-day timepoint, a lower score distribution was observed for the proxalutamide group than in the placebo group, **Figure 2A**. The median ordinal scale score was 1 (IQR 1–1) for proxalutamide versus 7 (IQR 2 to 8) for placebo. No difference was observed between sexes upon stratification for both placebo and proxalutamide at 28 days, **Figure 2B**. The overall 28-day recovery ratio was 1.81 (95% CI 1.60 to 2.00), which indicates that patients who took proxalutamide had an 81% higher recovery rate than those treated with placebo (95% CI 60 to 100), **Table 2**.

A high risk of all-cause mortality was observed for placebo (49.4%; 95% CI 44.0 to 54.7) compared to proxalutamide (11.0%; 95% CI 8.0 to 14.9). The number needed to treat (NNT) to prevent one death from COVID-19 in hospitalized patients over 28 days was 3 (95% CI 3 to 2). The risk ratio for death was 0.22 (95% CI 0.16 to 0.31), which indicates that treatment with proxalutamide reduced all-cause mortality rate over 28 days by 77.7%. Subgroup analysis by gender showed no interaction effects in any of the secondary outcome measures, **Table 2**. Other subgroup analyses of the 28-day recovery ratio and all-cause mortality risk by city of the study site are present in **Tables S3 and S4** of the **Supplementary Appendix**.

Figure 3 depicts the Kaplan-Meier survival curves and alive hospital discharge within 28 days for both the proxalutamide and placebo treated groups overall. The difference in the proportion surviving was evident as early as day 3 and increased over the remaining study period, which includes days after the 14-day therapy period (**Figure 3A**). This demonstrates there was no noticeable rebound effect if therapy was completed. The difference in the proportion of alive hospital discharge was statistically significant at day 2, and increased until day 11, reaching 75%, **Figure 3B**. Patients in the proxalutamide group during the study period were 84% (95% CI 89-76%) less likely to die than patients in the control group (hazard ratio for death 0.16 [95% CI, 0.11 to 0.24]). The median hospitalization time (days) was lower in the proxalutamide arm (8, IQR 6-13) compared to the placebo (12, IQR 8-18).

### Safety Outcomes

Adverse events observed during the course of the trial are detailed in **Table 3**. Adverse events grade 4 or 3 were more commonly observed in the placebo arm (40.9%) than in the proxalutamide arm (2.2%), *P*<0.001. Diarrhea was the only adverse event reported at higher proportions in those receiving proxalutamide (16.1%) versus those receiving placebo (3.3%), *P*=0.005. Irritability was reported in four patients treated with proxalutamide (1.3%) and none in the placebo group. Among males taking proxalutamide, four reported spontaneous erection (2.2%) versus none in males taking placebo.

**Table 3.**
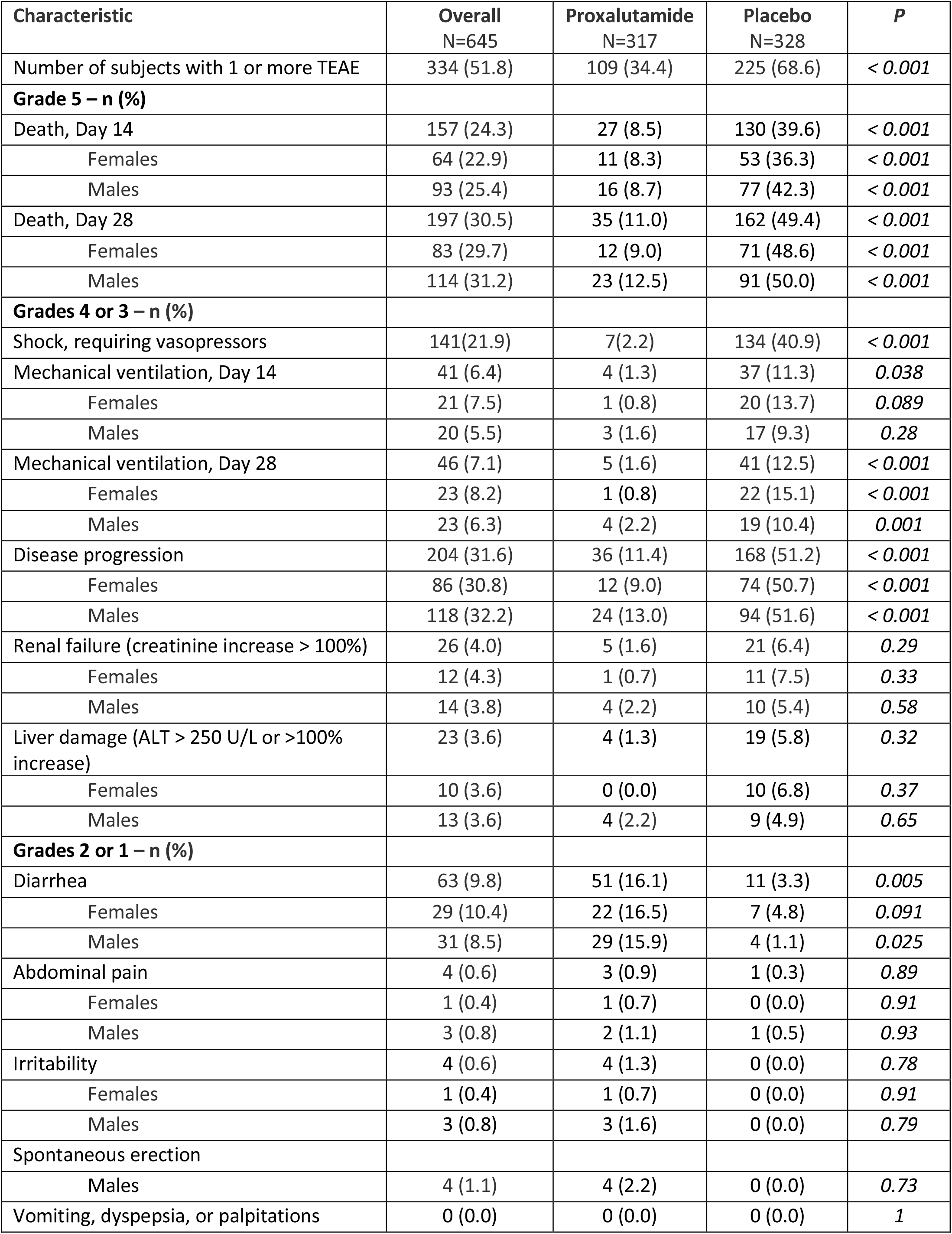
Treatment emergent adverse events (TEAE) in the patient population, per intention-to-treat analysis.

**Figure 3.**
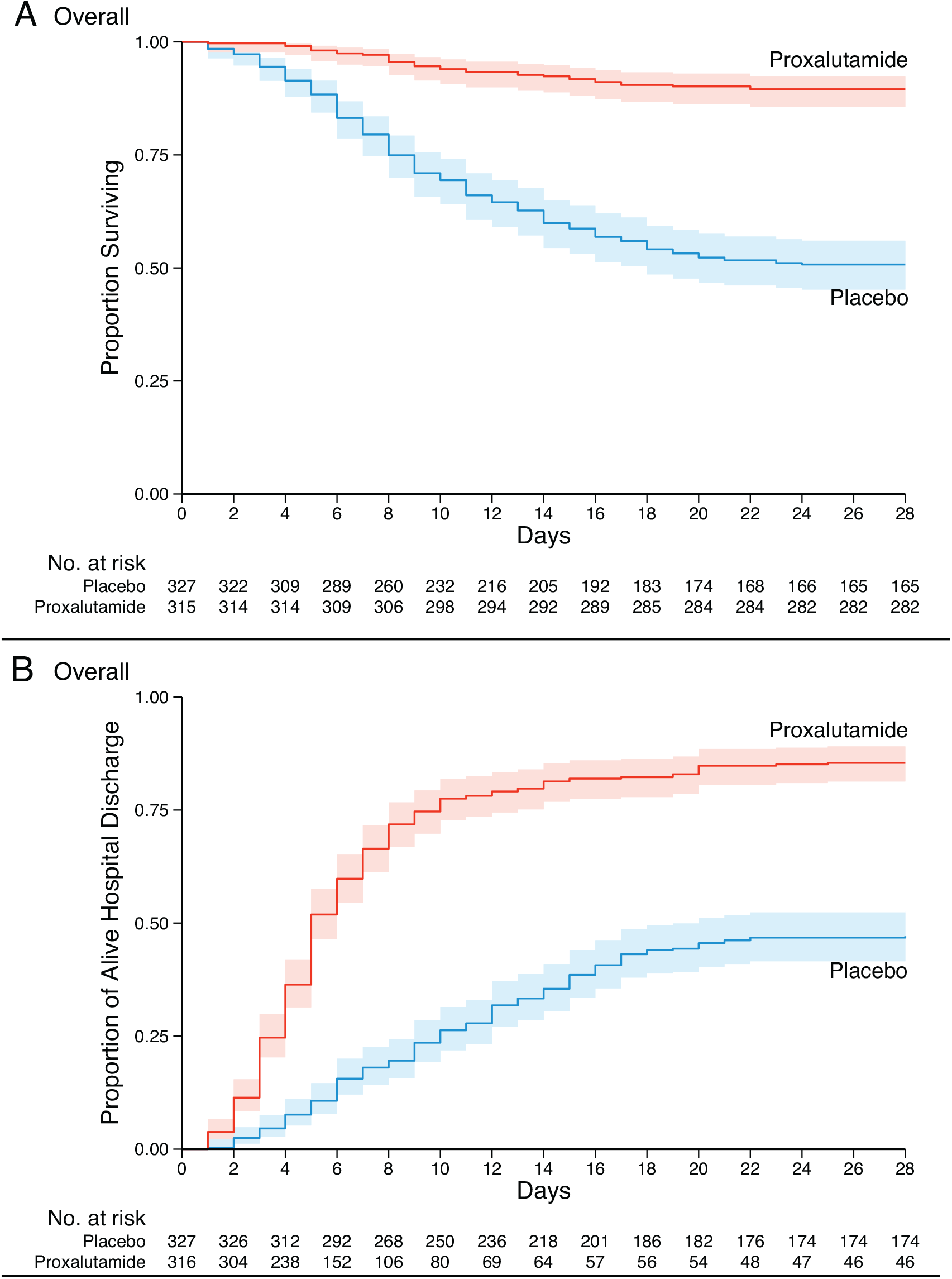
Kaplan–Meier estimates in the intention-to-treat analysis from randomization to day 28. Surviving (Panel A) and Alive Hospital Discharge (Panel B) for the overall population.

## Discussion

Here we demonstrate in a randomized, double-blind, placebo-controlled clinical trial that the use of proxalutamide, a second-generation nonsteroidal antiandrogen, reduced mortality, length of stay, and significantly improved clinical outcomes of hospitalized COVID-19 patients. These findings are consistent with our reports of the efficacy of proxalutamide in an outpatient setting,^10,15^ as well as reports using other antiandrogen regimens, including dutasteride and finasteride.^16,17^

Proxalutamide was a benefit to COVID-19 patients who were severely ill. At the time of the trial, the state of Amazonas experienced a surge in COVID-19 cases resulting in overcrowding at hospitals. As such, prioritized admission to hospitals resulted in admission of the most severely affected patients with COVID-19. Additionally, we found the mortality rate to be exceptionally high (49.4%) in the placebo group. Though the observed mortality rate may have been influenced by the emergent P.1 strain, the high mortality rate we observed in the placebo group is not unexpected based on past reports.^18,19^ The northern Amazonas region of Brazil have reported high mortality rates, ranging from 24-55% amongst hospitalized COVID-19 patients as early as April of 2020, while the average rates observed in all of Brazil range from 14-38%.^18,19^

As of March, 2021, the P.1 lineage is responsible for at least 70% of the current SARS-CoV-2 genomes sequenced in Brazil, and for at least 90% of the SARS-CoV-2 genomes in the state of Amazonas.^20^ In a post-hoc analysis, sequencing of viral genomes from patient samples obtained from the study sites and during the trial dates were found to be the P.1 lineage in all but one case (**Supplementary Appendix, Table S5**). P.1 is considered one of most relevant variants of concern (VOC) worldwide. This variant has demonstrated enhanced affinity for ACE-2 binding, potentially resulting in higher viral loads.^21^ In particular, the E484K substitution, previously demonstrated by our group as being positively selected in diverse lineages in Brazil,^22^ can induce immune escape with consequent reinfection and failure of other therapeutic modalities, such as convalescent plasma.^23^ It is encouraging to note, therefore, that proxalutamide was effective in a region with high P.1 prevalence.

Finding new or repurposed drugs active against VOCs and other SARS-CoV-2 harboring mutations is of paramount importance. Multiple agents have been explored in clinical trials as treatments for COVID-19. To date, the most promising of these treatments are remdesivir,^24^ a viral RNA-polymerase inhibitor, remdesivir plus the Janus kinase (JAK) inhibitor, baricitinib,^25^ and dexamethasone,^26^ a glucocorticoid without mineralocorticoid effect. Remdesivir has been reported to reduce the median time to recovery of hospitalized COVID-19 patients to 10 days, down from 15 days observed with placebo.^24^ The mortality rate of patients treated with remdesivir was 11.4% at 29 days compared to 15.2% in the placebo group, though not reaching statistical significance (HR 0.73; 95% CI, 0.52 to 1.03).^24^ Similarly, the combination of remdesivir and baricitinib trended to a reduction in 28-day mortality from 7.8% to 5.1% (HR 0.65; 95% CI, 0.39 to 1.09).^25^ It is difficult to compare results across trials, but we observed the clinical benefits of proxalutamide to be superior to either of these treatments with greater than 77.7% reduction in 28-day mortality rate under ITT analysis (HR of 0.16; 95% CI, 0.11 to 0.24). Direct comparisons among therapies are warranted to delineate effective regimen(s). Dexamethasone has also been demonstrated to benefit the most severe COVID-19 patients; among those patients requiring mechanical ventilation, dexamethasone was shown to reduce mortality compared to placebo (29.3% vs. 41.4%; rate ratio, 0.64; 95% CI, 0.51 to 0.81).^26^ All of the participants in this trial received corticosteroids, but there was still a survival advantage to receiving proxalutamide.

Treatment emergent adverse events associated with proxalutamide were limited to diarrhea. Diarrhea was also reported to be more frequent among the proxalutamide group in an outpatient trial.^11^ In contrast, severe adverse events of renal failure and hepatic damage were associated with placebo, **Table 3**. We interpret this finding as the natural progression of COVID-19 in the placebo group, which further supports consideration of proxalutamide as a therapy for hospitalized patients. While Phase 1 safety data for proxalutamide supports its use for 28 days in both men and women (**Protocol for details**), it has been studied primarily in prostate cancer patients, therefore, long term side effects should be assessed in future studies. Alternatively, other approved molecules of the same class may show similar results, such as apalutamide, enzalutamide, darolutamide, bicalutamide, or flutamide.

Important limitations are present in this study. First, the remote locations of many of the study sites created operational difficulties that led to an unbalanced distribution of proxalutamide and placebo amongst sites (details provided in **Supplementary Appendix**). Minor differences in effects of proxalutamide between sites may have resulted from factors such as local infrastructure and age distribution (**Figures S5 and S6**). However, the possibility of effect sizes skewing in favor of the proxalutamide group due to its disproportional concentration in the smaller hospitals is unlikely because a similar size effect was observed in subgroup analysis of all hospital sites (**Tables S3 and S4**). Second, the severity of patients admitted to Amazonas hospitals during the trial did not allow us to test proxalutamide in many hospitalized patients who did not require supplemental oxygen (ordinal score 3). However, we have previously shown that proxalutamide reduced hospitalizations and improved symptom recovery and viral clearance in outpatients with mild-moderate disease.^10^ Moreover, the subgroup analysis showed patients with baseline scores of 3-5 benefited from proxalutamide. Third, more patients used colchicine in the placebo arm despite randomization. This most likely reflects additional therapeutic interventions associated with longer hospitalization in the placebo group. Colchicine is unlikely, however, to have contributed to increased mortality directly as there is either no effect,^27^ with 28-day mortality rate ratio of 1.01 in the RECOVERY platform trial (95% CI, 0.93 to 1.10)^28^, or limited benefit in COVID-19.^29–31^ Forth, remdesivir was not available to our patients. Despite the lack of a direct acting antiviral, however, patients receiving proxalutamide had a substantial benefit. Lastly, a dropout rate of approximately 9-11% was observed in both study arms. The explanation for this participant attrition varied and was seen in both arms, nevertheless, proxalutamide retained its efficacy in the ITT analysis.

This trial has generated important questions requiring further study. Despite being an antiandrogen therapy, women treated with proxalutamide responded similarly to proxalutamide-treated men (**Fig 2 and Table 2**). Additionally, the significant improvement observed in severely-ill patients appeared disproportionate to the proposed mechanism of reducing viral entry **(Table 2, Baseline score 6)**. These findings suggest additional mechanism(s) of action beyond androgen antagonism and TMPRSS2 knockdown that will require additional investigation. Because of the logistical difficulties executing this trial and the high mortality in Brazil during the study period, our findings must be evaluated in other populations. It is urgent that additional trials examine non-steroidal antiandrogens in different settings.

## Conclusions

Hospitalized COVID-19 patients receiving treatment with proxalutamide had a 128% higher recovery rate than those treated with placebo at day 14. All-cause mortality was reduced by 77.7% over 28 days. Further studies of proxalutamide and other antiandrogen therapies in COVID-19 patients in different settings and locations are urgently needed.

## Data Availability

Dataset is available under request and specific justification.

## Supplementary Appendix

### Additional Clinical Trial Sites Details

12 sites were approved to recruit patients. However, recruitment of patients was rapid and 4 sites were not able to participate before the trial had reached recruitment goals.

Sites that recruited patients during the trial and number of patients randomized (n):

1. Hospital Samel, Manaus, Amazonas, Brazil (99)
2. Hospital Oscar Nicolau, Manaus, Amazonas, Brazil (108)
3. Hospital Prontocord, Manaus, Amazonas, Brazil (188)
4. Hospital Regional José Mendes, Itacoatiara, Amazonas, Brazil (112)
5. Hospital Raimunda Francisca Dinelli da Silva, Maues, Amazonas, Brazil (12)
6. Hospital Regional Dr. Hamilton Maia Cidae, Manicore, Amazonas, Brazil (5)
7. Hospital Regional Jofre Cohen, Parintins, Amazonas, Brazil (103)
8. Hospital de Campanha de Manacapuru, Manacapuru, Amazonas, Brazil (18)

**Manaus, Amazonas, Brazil: 395 patients randomized**

**Other cities, Amazonas, Brazil: 250 patients randomized**

Sites approved to participate in the study that did not recruit patients before trial closed enrollment (no patients randomized):

1. Hospital Regional de Coari Prof. Dr. Odair Carlos Geraldo, Coari, Amazonas, Brazil
2. Hospital de Campanha de Barcelos, Barcelos, Amazonas, Brazil
3. Hospital Regional de Labrea, Labrea, Amazonas, Brazil
4. Hospital Regional de Humaitá, Humaitá, Amazonas, Brazil

## Additional Inclusion and Exclusion Criteria Details

### Inclusion Criteria

- Subjects enrolled in this study were required to meet the following key acceptance criteria:
- Admitted to the hospital with symptoms of COVID-19
- Male and females age ≥18 years old
- Laboratory confirmed positive SARS-CoV-2 rtPCR test within 7 days prior to randomization
- Clinical status on the COVID-19 Ordinal Scale of 3, 4, 5, or 6
- Coagulation: INR ≤ 1.5×ULN, and APTT ≤ 1.5×ULN
- Subject (or legally authorized representative) gives written informed consent prior to performing any study procedures
- Subject (or legally authorized representative) agree that subject will not participate in another COVID-19 trial while participating in this study

### Exclusion Criteria

Subjects were not to be enrolled into the study if it was determined upon pre-study examination that they met the following key criteria:

- Subject enrolled in a study to investigate a treatment for COVID-19
- Requires mechanical ventilation
- Subject taking an anti-androgen of any type including: androgen depravation therapy, 5-alpha reductase inhibitors, etc.
- Patients who are allergic to the investigational product or similar drugs (or any excipients);
- Subjects who have malignant tumors in the past 5 years, with the exception of completed resected basal cell and squamous cell skin cancer and completely resected carcinoma in situ of any type
- Subjects with known serious cardiovascular diseases, congenital long QT syndrome, torsade de pointes, myocardial infarction in the past 6 months, or arterial thrombosis, or unstable angina pectoris, or congestive heart failure which is classified as New York Heart Association (NYHA) class 3 or higher, or left ventricular ejection fraction (LVEF) < 50%, QTcF > 450 ms
- Subjects with uncontrolled medical conditions that could compromise participation in the study (e.g., uncontrolled hypertension, hypothyroidism, diabetes mellitus)
- Known diagnosis of human immunodeficiency virus (HIV), hepatitis C, active hepatitis B, treponema pallidum (testing is not mandatory)
- Alanine Transaminase (ALT) or Aspartate Transaminase (AST) > 5 times the upper limit of normal.
- Estimated glomerular filtration rate (eGFR) < 30 ml/min
- Severe kidney disease requiring dialysis
- Women of child-bearing potential, defined as all women physiologically capable of becoming pregnant, unless they are using highly effective contraception, as shown below, throughout the study and for 3 months after stopping GT0918 treatment. Highly effective contraception methods include:

1. Total Abstinence (when this is in line with the preferred and usual lifestyle of the patient. Periodic abstinence (e.g., calendar, ovulation, symptothermal, post-ovulation methods) and withdrawal are not acceptable methods of contraception, or
2. Use of one of the following combinations (a+b or a+c or b+c):
  a. Use of oral, injected or implanted hormonal methods of contraception or other forms of hormonal contraception that have comparable efficacy (failure rate < 1%), for example hormone vaginal ring or transdermal hormone contraception.
  b. Placement of an intrauterine device (IUD) or intrauterine system (IUS); c: Barrier methods of contraception: Condom or Occlusive cap (diaphragm or cervical/vault caps) with spermicidal foam/gel/film/cream/vaginal suppository;
3. Female sterilization (have had prior surgical bilateral oophorectomy with or without hysterectomy) or tubal ligation at least six weeks before taking study treatment. In case of oophorectomy alone, only when the reproductive status of the woman has been confirmed by follow-up hormone level assessment;
4. Male sterilization (at least 6 months prior to screening). For female patients on the study, the vasectomized male partner should be the sole partner for that patient;
5. In case of use of oral contraception women should have been stable for a minimum of 3 months before taking study treatment. Women are considered post-menopausal and not of child bearing potential if they have had 12 months of natural (spontaneous) amenorrhea with an appropriate clinical profile (e.g., age appropriate, history of vasomotor symptoms) or have had surgical bilateral oophorectomy (with or without hysterectomy) or tubal ligation at least six weeks ago. In the case of oophorectomy alone, only when the reproductive status of the woman has been confirmed by follow up hormone level assessment, is she considered not of child bearing potential.

- Sexually active males must use a condom during intercourse while taking the drug and for 3 months after stopping treatment and should not father a child in this period. A condom is required to be used also by vasectomized men in order to prevent delivery of the drug via seminal fluid
- Subject likely to transfer to another hospital within the next 28 days
- Subject (or legally authorized representative) not willing or unable to provide informed consent

### Additional Randomization Procedures

Before the onset of the trial, a randomization table was created using a web-based randomization software (sealedenvelope.com/simple-randomiser/v1/lists) using 4, 6 and 8 block sizes and a list length for 662 treatment packages of identical appearance of either active or placebo group. A pharmacist sealed and labeled each package with a 3-character random-generated code. The packages contained a total of 6 blister packs of 7 individually sealed tablets, which accounted for a total of 42 tablets per treatment. The package instructions stated: take 3 tablets by mouth once a day for 14 consecutive days. The study was double-blinded with the identification of the group assignment known by the study monitor and the pharmacist who labeled the packages, who did not participate in dispensing the packages. Patients who were discharged before treatment day 14 had the remaining tablets dispensed as to complete the full 14-day treatment course, and were actively evaluated for compliance daily until day 14. All centers followed the same protocol.

At the three hospital sites in Manaus, the treatments were dispensed by the hospital pharmacy at random. Due to logistic difficulties in smaller remote hospitals with limited resources, and the possibility of unmonitored sharing of identical blister packs between patients, a procedure was undertaken to ensure non-sharing within the same site: boxes containing 5-50 treatment packages of the same blinded randomized treatment group were sealed for delivery to remote sites. Each box was delivered by a blinded research assistant directly to the research team at each remote site according to the local demand and enrollment capabilities. A new randomized box was delivered to the remote sites only after dispensing the previous box. The remote sites were not informed that each box contained a single arm, and were instructed to follow the protocol and register in the case report forms the individual package 3-character code dispensed for each patient. This operation procedure introduced a bias in the distribution of drug and placebo between sites. **Table S1** details the drug and placebo distribution per site and the randomization scheme used. **Figure S1** shows the enrollment over the time of the study. We do not believe the drug and placebo distribution bias impacted the overall results of the study as more proxalutamide was dispensed to the remote hospitals with less resources.

**Table S1.**
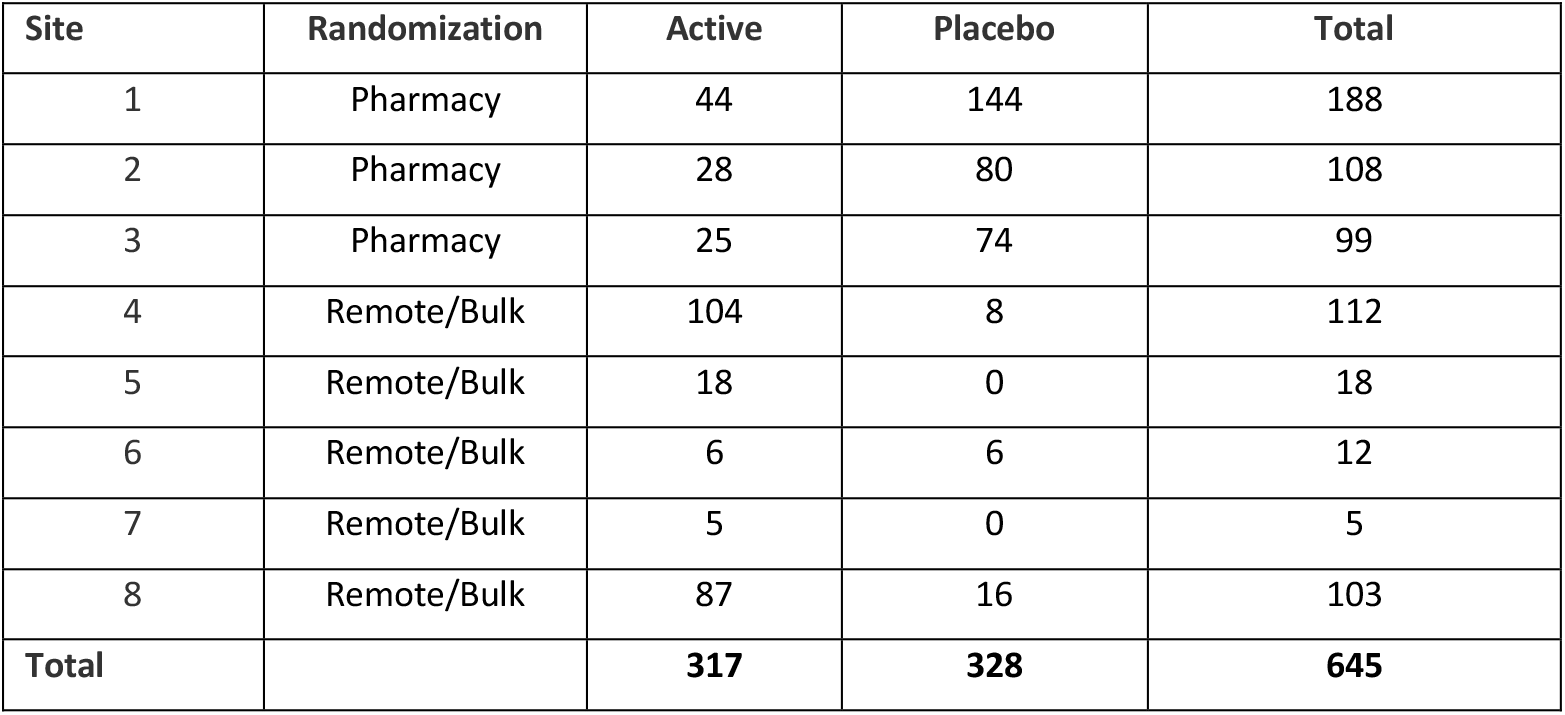
Group Distribution per Site.

### Interim Analysis and Public Disclosure, March 10^th^ 2021

An interim analysis was planned per protocol at 50% recruitment of the target sample size. Owing to the surge in cases experienced during the trial, however, patient recruitment was rapid and exceeded 50% before the 28-day safety assessment. Additionally, when 50% recruitment goal was achieved (Feb 14) no endpoint data was available. Accordingly, the interim analysis was performed later, in March. The preliminary results were presented to the public on March 10, 2021, in accordance with Brazilian National Research Ethics Committee guidance (National Health Council Resolution Number 466, III.2.m). At the time of the interim analysis, the majority of patients, 99%, had been randomized; only 9 additional patients were randomized after the event. **Figure S1** details the number of patients randomized to study before and after March 10th. It is noteworthy, that only 55 of the 645 patients had yet to complete the 14-day course of study intervention at the time of the public disclosure. As such, we do not believe the disclosure impacted recruitment, data collection, or interpretation of results.

**Figure S1.**
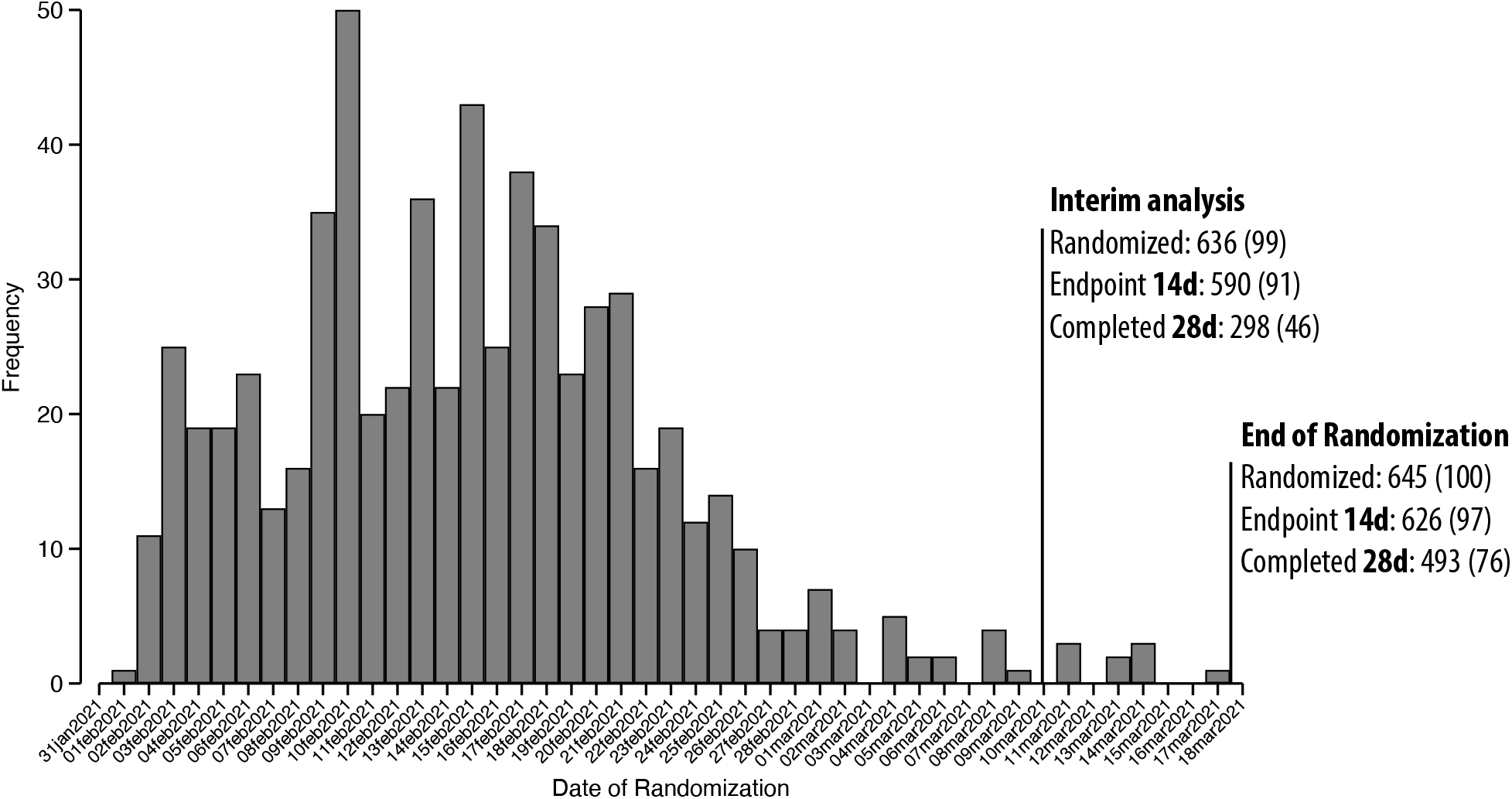
Randomization/recruitment timeline.

### Dosage Administration and Compliance Procedures

Proxalutamide 300 mg (3 × 100 mg tablets) or matching placebo was taken orally once daily with or without food, therapy was initiated soon after randomization. Treatment compliance was monitored and recorded while the patient was hospitalized. An accurate and current accounting of the dispensing of the study drug for each subject was maintained on an ongoing basis by the Investigator or delegated personnel. The number of study drug tablets dispensed to the subject was recorded on the Investigational Product Accountability Log. Patients who were discharged before treatment day 14 had the remaining tablets dispensed as to complete the full 14-day treatment course and were actively evaluated for compliance daily until day 14. All centers followed the same protocol.

### Baseline COVID-19 8-point Ordinal Scale

The protocol was amended before initiation of the trial because of prevailing disease in the participating hospitals. In the original protocol, the inclusion was limited to COVID-19 8-point ordinal scale scores 3, 4, and 5, i.e., did not include patients requiring non-invasive ventilation or high flow oxygen devices.

Prior to the start of the trial, Brazil, and particularly the state of Amazonas, experienced a surge in COVID-19 resulting in an increase in severity of cases. The majority of hospitalized patients on the days preceding the trial were on high flow oxygen devices (COVID-19 8-point ordinal scale score 6). As such, the decision was made to include score 6 on the day of the initiation of recruitment. All patients, including COVID-19 8-point ordinal scale score 6, were randomized in the same 1:1 ratio as indicated in the **Protocol**.

We did not perform randomization stratified per site as a result of batch delivery of drugs or placebo (described above). This led to an unbalance between study arms within hospital sites. In three small hospital centers the randomization strategy had to be modified due to: 1) logistic difficulties in taking the drugs to these remote hospitals and 2) limited financial resources. This was done to avoid unblinding and unmonitored sharing of blister packs among hospitals and patients. We are aware that these changes in the randomization scheme increased the risk of bias in the randomization procedure. However, in the 3 sites located in the large urban city of Manaus with higher hospital certification standards, where randomization could be done as originally planned, a lower mortality rate in both arms was observed, as well as a lower mortality risk ratio (**Table S4**).

**Table S2.**
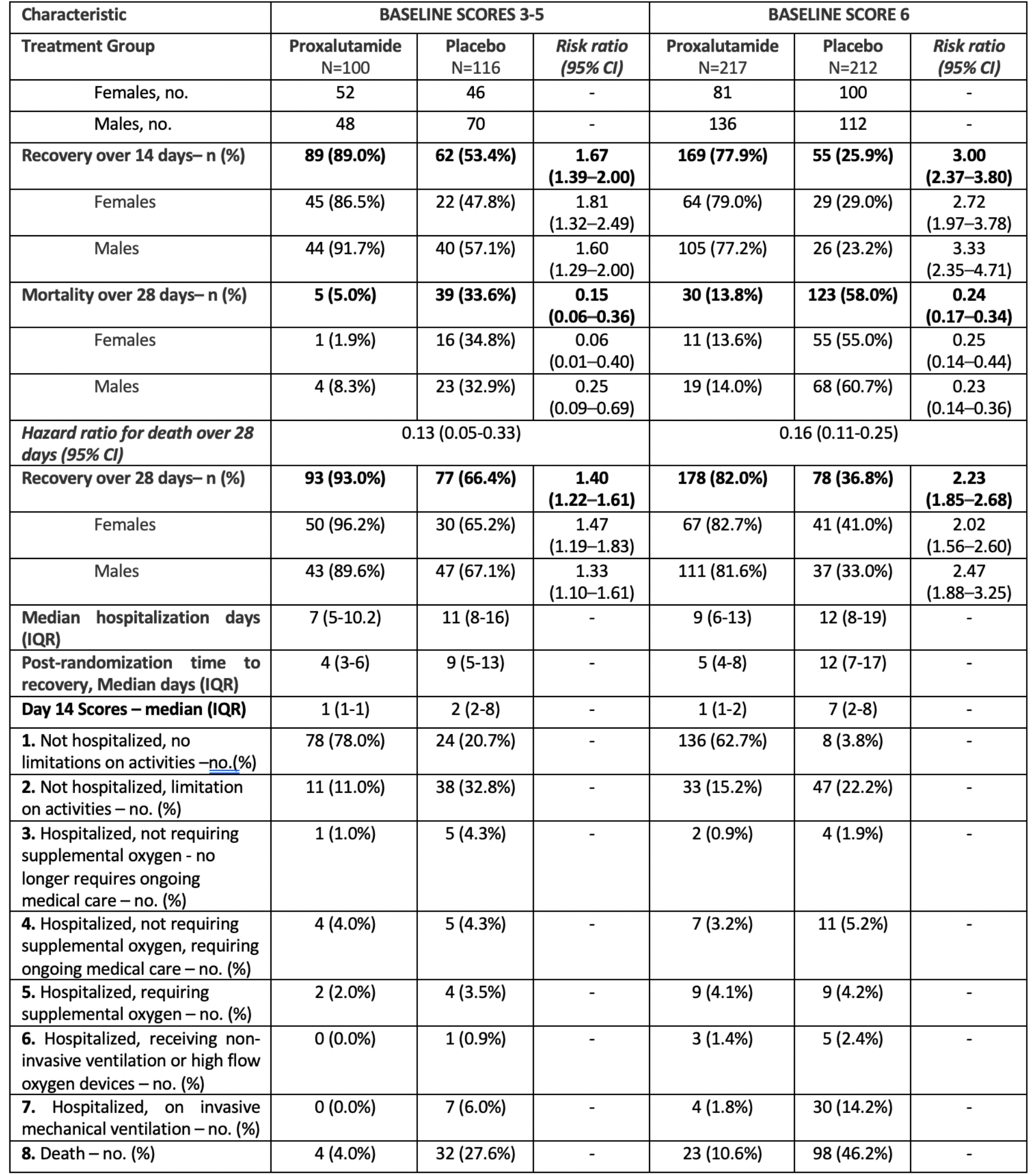
Coronavirus disease 2019 8-point ordinal scale scores distribution and outcomes by baseline scores.

**Table S3.**
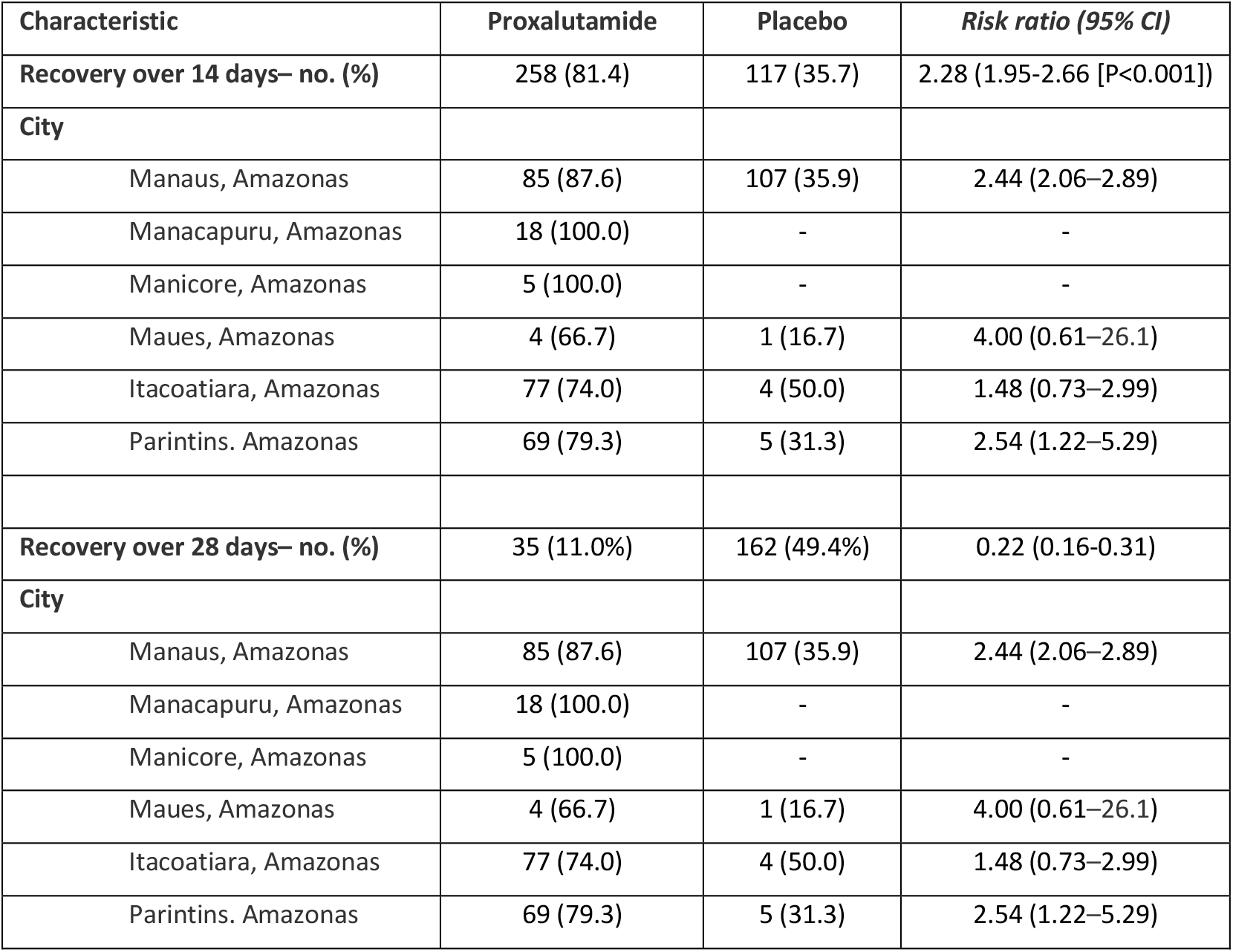
Recovery (coronavirus disease 2019 8-point ordinal scale scores 1 or 2, alive hospital discharge) over 14- and 28-days post-randomization stratified by city.

**Table S4.**
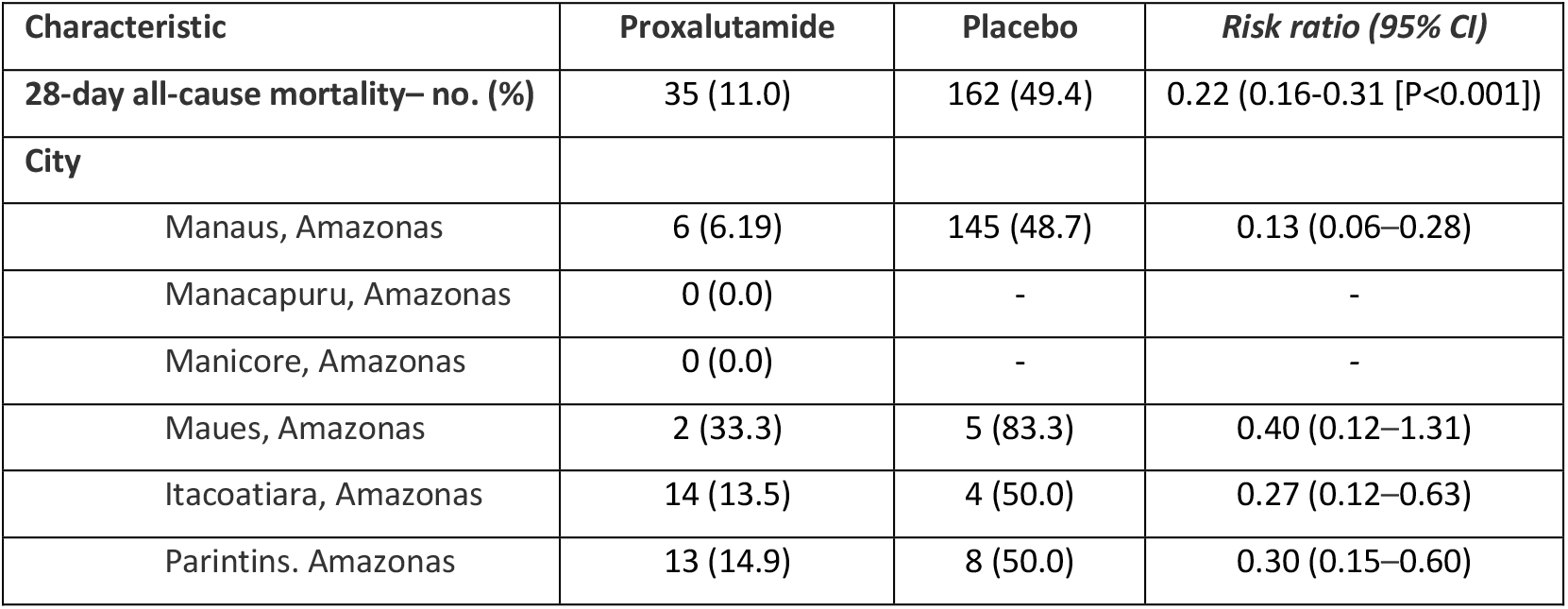
All-cause mortality over 28 days post-randomization stratified by city.

**Figure S2.**
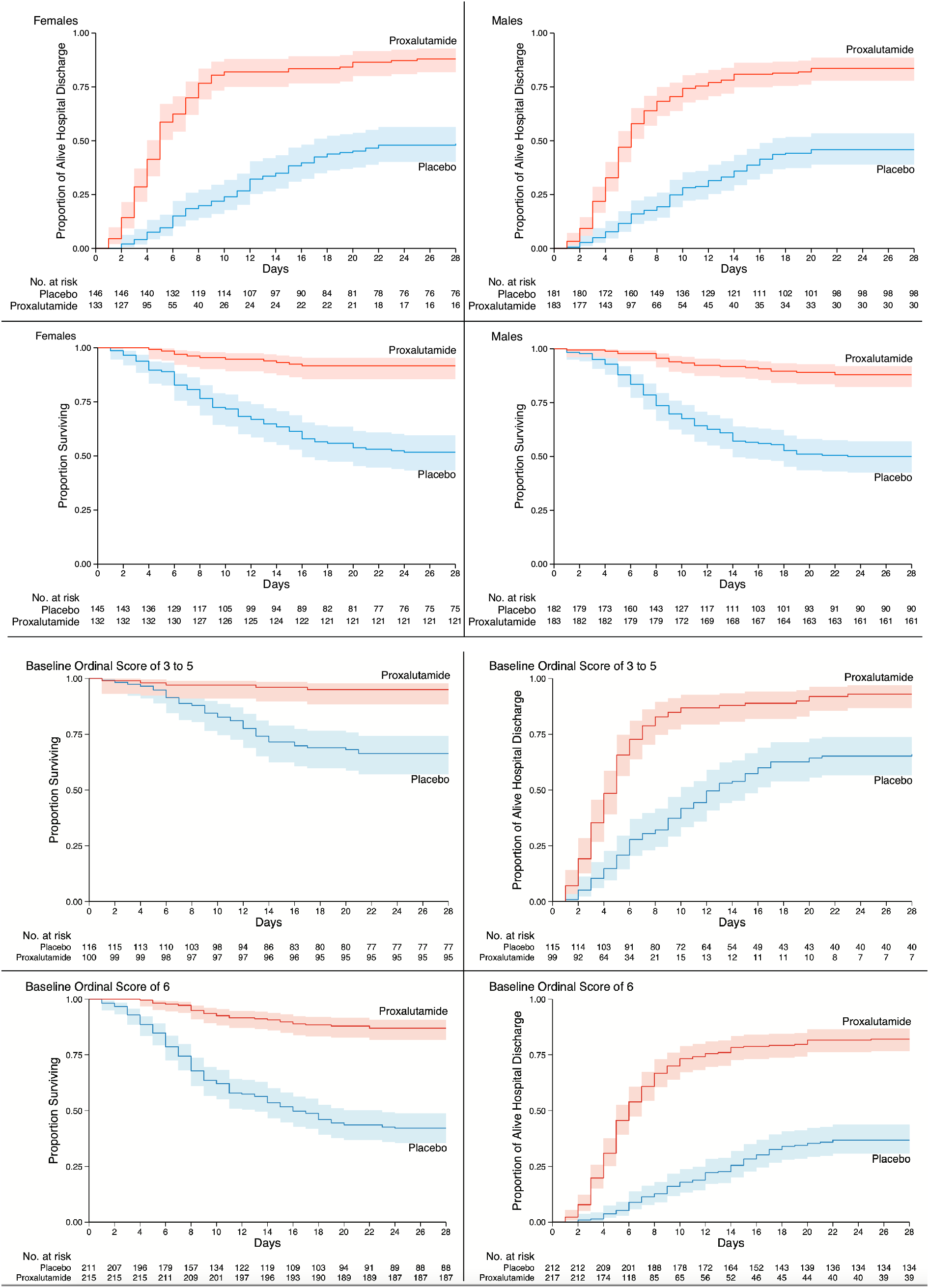
Kaplan–Meier estimates from randomization to Day 28. Alive Hospital Discharge and Proportion Surviving by sex and baseline ordinal scale.

**Figure S3.**
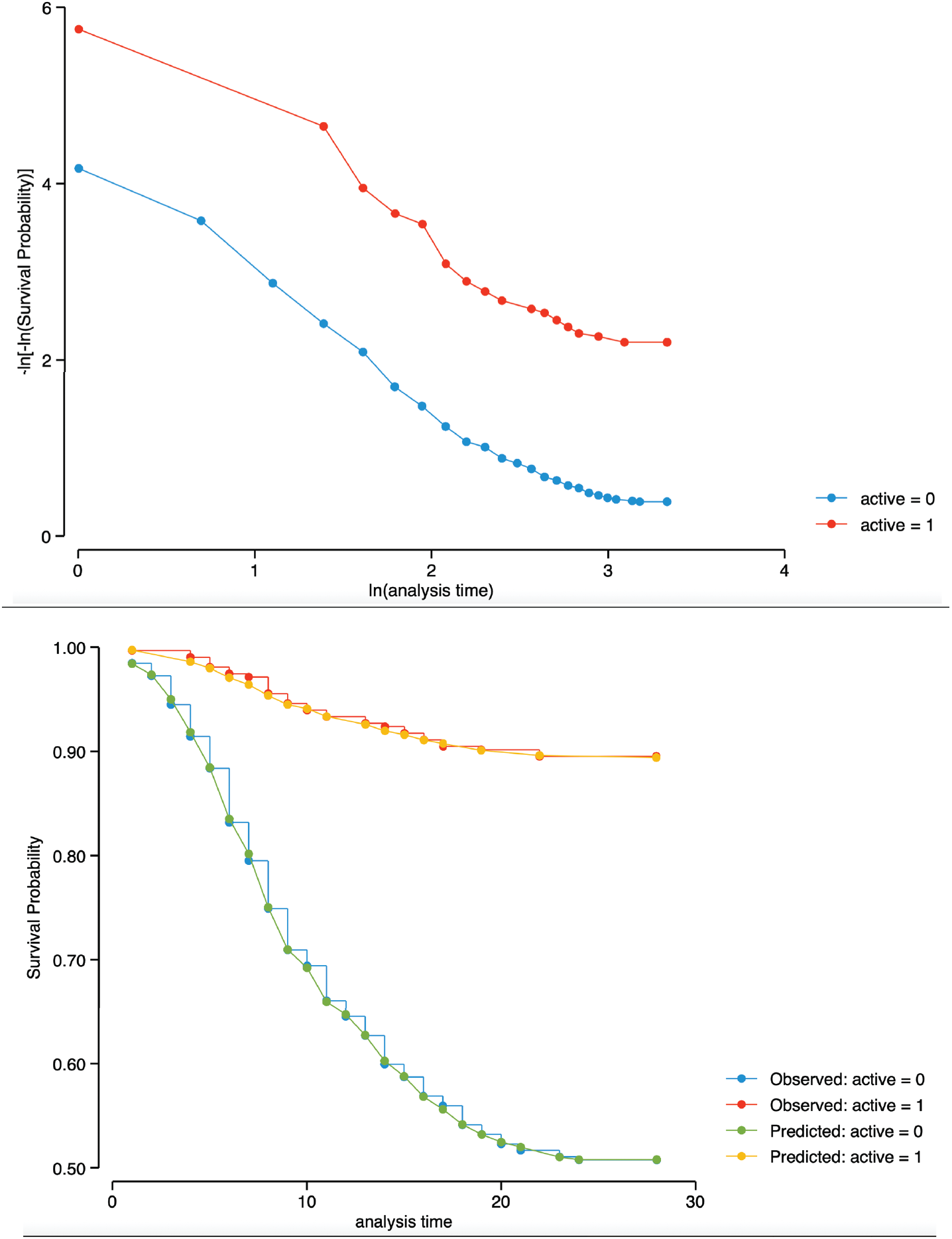
Graphical assessment of proportional-hazards assumption (hazard ratio over 28 days post-randomization). **Top:** figure displays lines that are parallel, implying that the proportional-hazards assumption for the therapy groups has not been violated. **Bottom:** This is confirmed by Kaplan-Meier versus predicted survival, where the observed values and predicted values are close together for the therapy groups. Proportional-hazards assumption on the basis of Schoenfeld residuals revealed a global test with P value = 0.7986, therefore, we can assume the proportional hazards.

### SARS-CoV-2 Lineage Determination

For this post hoc analysis, clinical samples from patients admitted to one of the participating centers testing positive for SARS-CoV-2 in a first RT-qPCR had their samples submitted to a second RT-qPCR performed by BiomeHub (Florianópolis, Santa Catarina, Brazil), using charite-berlin protocol. Only samples with quantification cycle (Cq) below 30 for at least one primer were processed for SARS-CoV-2 genome sequencing by the BiomeHub laboratory.

To perform the SARS-CoV-2 genome sequencing, total RNAs were prepared as in the reference protocol (dx.doi.org/10.17504/protocols.io.befyjbpw) using SuperScript IV (Invitrogen) for cDNA synthesis and Platinum Taq High Fidelity (Invitrogen) for specific viral amplicons. The cDNA obtained were subsequently used for the library preparation with Nextera Flex (Illumina) and quantified with Picogreen and Collibri Library Quantification Kit (Invitrogen). The sequencing was performed on MiSeq 150×150 runs with 500xSARS-CoV-2 coverage (50-100 mil reads/per sample).

The SARS-CoV-2 genome assembly was generated by an in-house pipeline from BiomeHub (Florianópolis, Santa Catarina, Brazil). The remotion of adapters and read trimming in 150 nt were performed with fastqtools.py, followed by the reads mapping to the reference SARS-CoV-2 genome (GenBank accession number NC_045512.2) with Bowtie v2.4.2 (additional parameters: end-to-end and very-sensitive). The mapping coverage and sequencing depth were obtained with samtools v1.11 (minimum base quality per base (Q) ≥ 30). Consensus genome sequences were then generated with bcftools mpileup (Q ≥ 30; depth (d) ≤ 1,000) combined with bcftools filter (DP>50) and bcftools consensus v1.11. Finally, the identification of the SARS-CoV-2 virus lineages was performed by the Pangolin v2.3.8 web server (https://github.com/cov-lineages/pangolin).

**Table S5.**
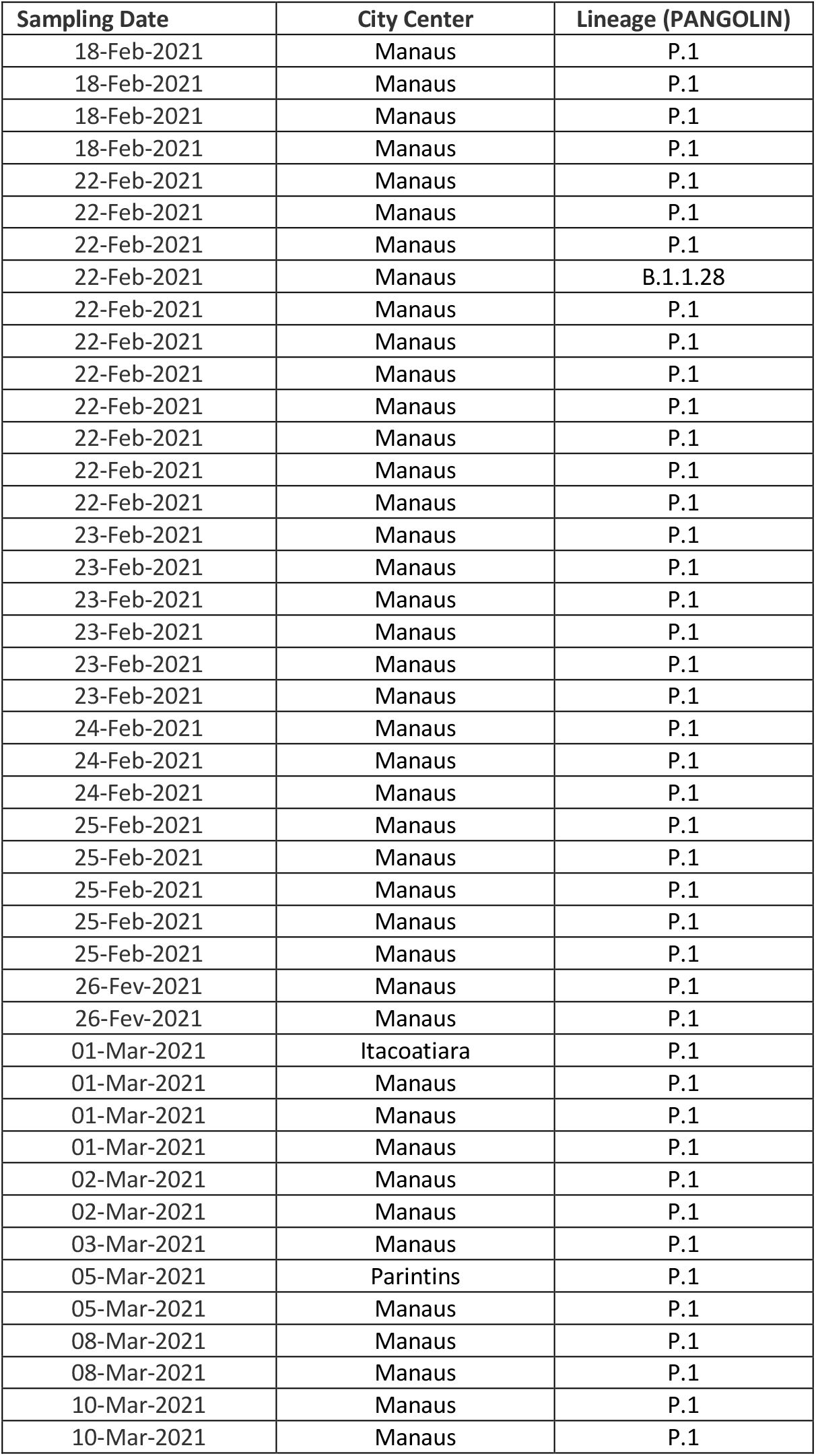
Sequencing of SARS-CoV-2 in 44 Randomly Selected Patients.

**Figure S4.**
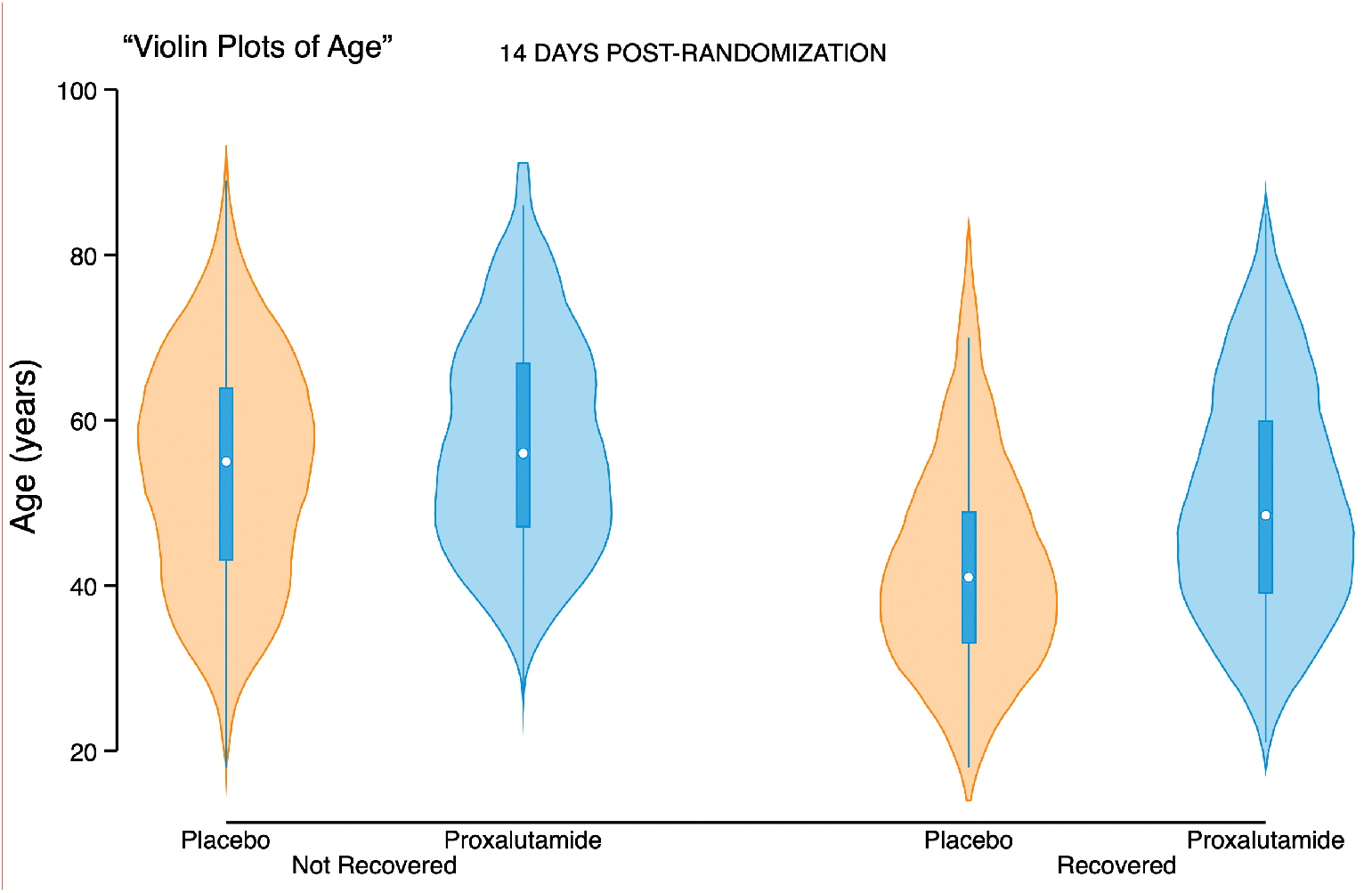
Violin Plots for Age, for alive hospital discharge over the first 14 days post-randomization, per treatment group.

**Figure S5.**
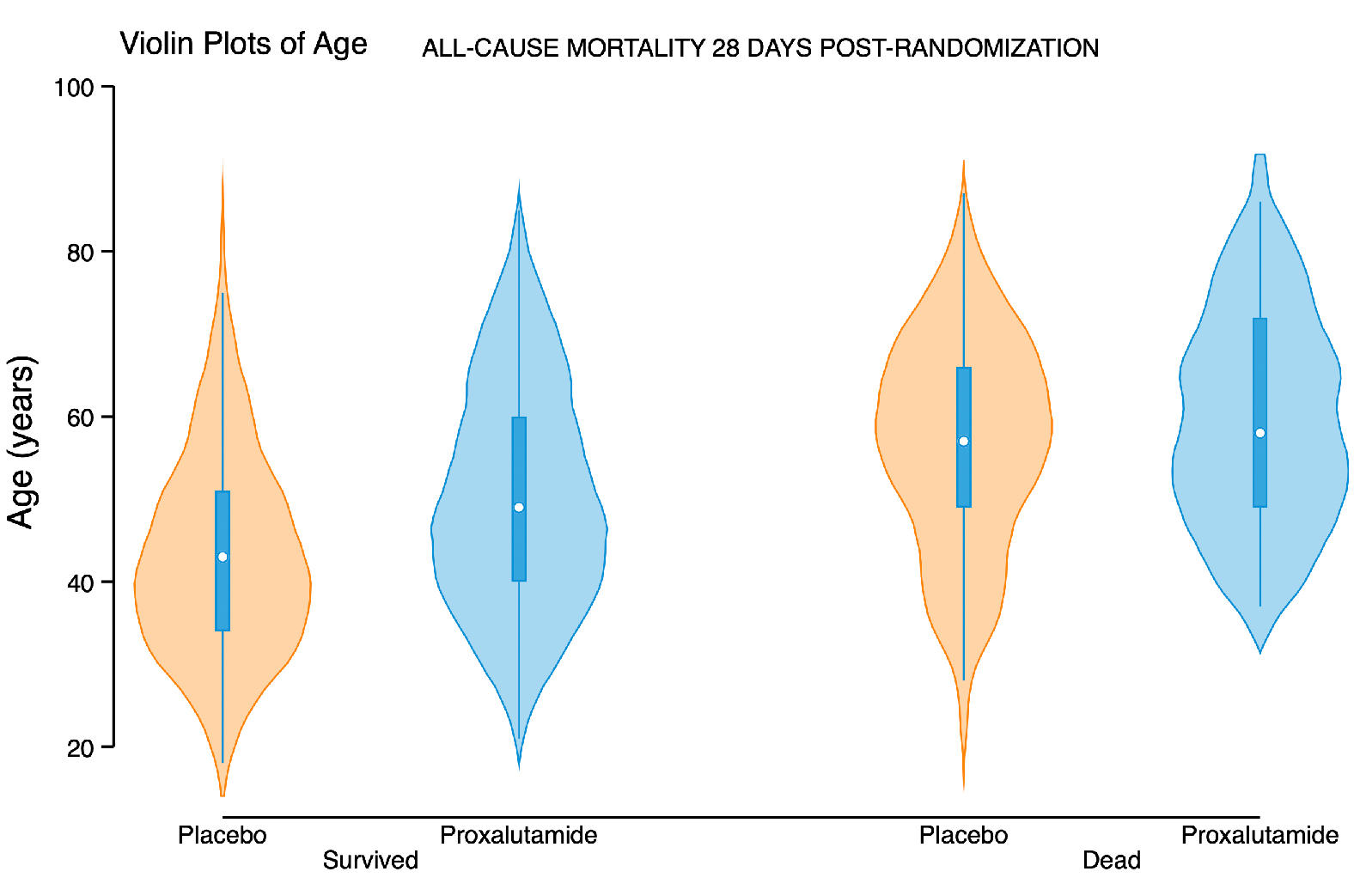
Violin Plots for Age, for all-cause mortality over the 28 days post-randomization, per treatment group.

**Figure S6.**
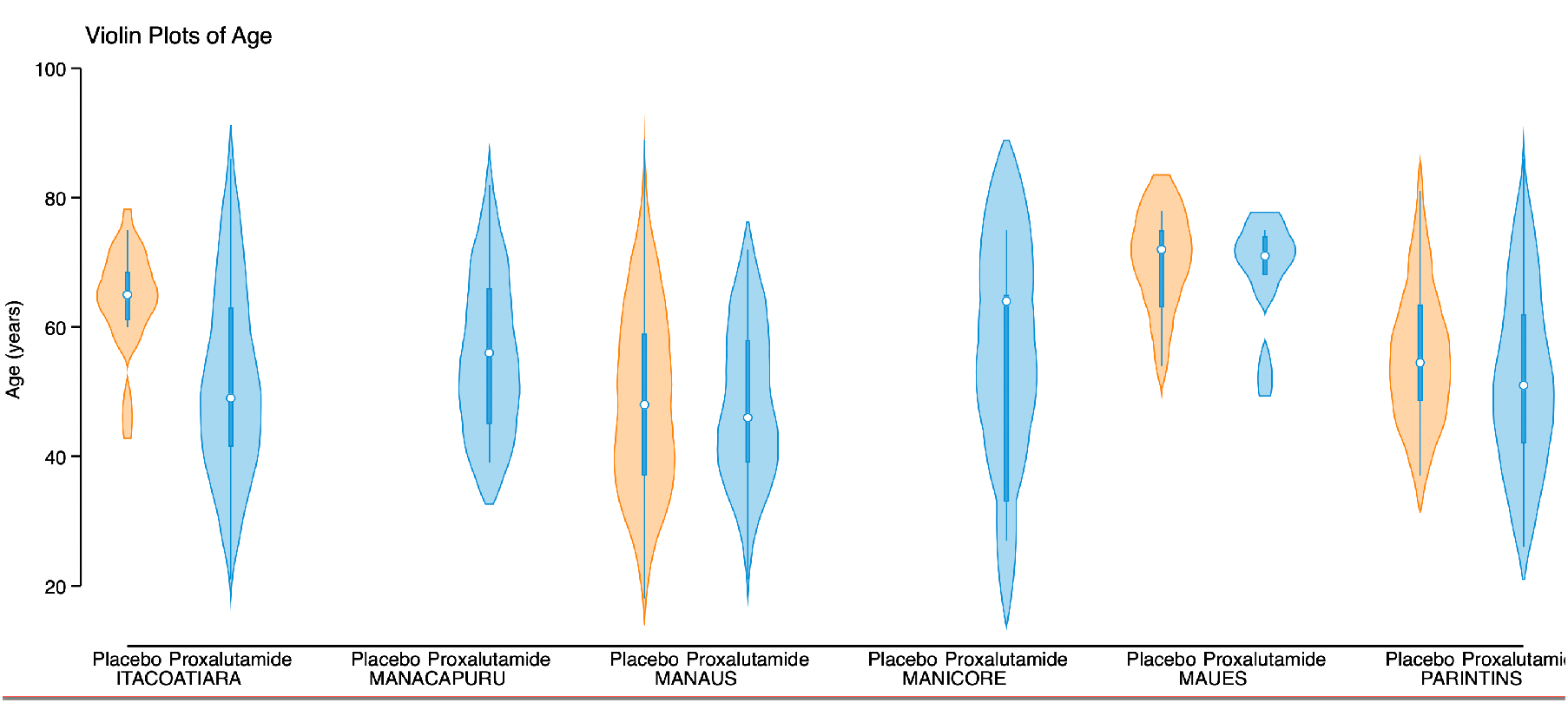
Violin Plots for Age, for cities, per treatment group.

