## Supplementary Appendix for "Efficacy of Proxalutamide in Hospitalized COVID-19 Patients: A Randomized, Double-Blind, Placebo-Controlled, Parallel-Design Clinical Trial"

**Additional Clinical Trial Sites Details**

12 sites were approved to recruit patients. However, recruitment of patients was rapid and 4 sites were not able to participate before the trial had reached recruitment goals.

Sites that recruited patients during the trial and number of patients randomized (n):

1. Hospital Samel, Manaus, Amazonas, Brazil (99)
2. Hospital Oscar Nicolau, Manaus, Amazonas, Brazil (108)
3. Hospital Prontocord, Manaus, Amazonas, Brazil (188)
4. Hospital Regional José Mendes, Itacoatiara, Amazonas, Brazil (112)
5. Hospital Raimunda Francisca Dinelli da Silva, Maues, Amazonas, Brazil (12)
6. Hospital Regional Dr. Hamilton Maia Cidae, Manicore, Amazonas, Brazil (5)
7. Hospital Regional Jofre Cohen, Parintins, Amazonas, Brazil (103)
8. Hospital de Campanha de Manacapuru, Manacapuru, Amazonas, Brazil (18)

**Manaus, Amazonas, Brazil: 395 patients randomized**
**Other cities, Amazonas, Brazil: 250 patients randomized**

Sites approved to participate in the study that did not recruit patients before trial closed enrollment (no patients randomized):

9. Hospital Regional de Coari Prof. Dr. Odair Carlos Geraldo, Coari, Amazonas, Brazil
10. Hospital de Campanha de Barcelos, Barcelos, Amazonas, Brazil
11. Hospital Regional de Labrea, Labrea, Amazonas, Brazil
12. Hospital Regional de Humaitá, Humaitá, Amazonas, Brazil

#### Table S1. Group Distribution per Site

| Site | Randomization | Active | Placebo | Total |
|---|---|---|---|---|
| 1 | Pharmacy | 44 | 144 | 188 |
| 2 | Pharmacy | 28 | 80 | 108 |
| 3 | Pharmacy | 25 | 74 | 99 |
| 4 | Remote/Bulk | 104 | 8 | 112 |
| 5 | Remote/Bulk | 18 | 0 | 18 |
| 6 | Remote/Bulk | 6 | 6 | 12 |
| 7 | Remote/Bulk | 5 | 0 | 5 |
| 8 | Remote/Bulk | 87 | 16 | 103 |
| **Total** | | **317** | **328** | **645** |

#### Interim Analysis and Public Disclosure, March 10th 2021

An interim analysis was planned per protocol at 50% recruitment of the target sample size. Owing to the surge in cases experienced during the trial, however, patient recruitment was rapid and exceeded 50% before the 28-day safety assessment. Additionally, when 50% recruitment goal was achieved (Feb 14) no endpoint data was available. Accordingly, the interim analysis was performed later, in March. The preliminary results were presented to the public on March 10, 2021, in accordance with Brazilian National Research Ethics Committee guidance (National Health Council Resolution Number 466, III.2.m). At the time of the interim analysis, the majority of patients, 99%, had been randomized; only 9 additional patients were randomized after the event. **Figure S1** details the number of patients randomized to study before and after March 10th. It is noteworthy, that only 55 of the 645 patients had yet to complete the 14-day course of study intervention at the time of the public disclosure. As such, we do not believe the disclosure impacted recruitment, data collection, or interpretation of results.

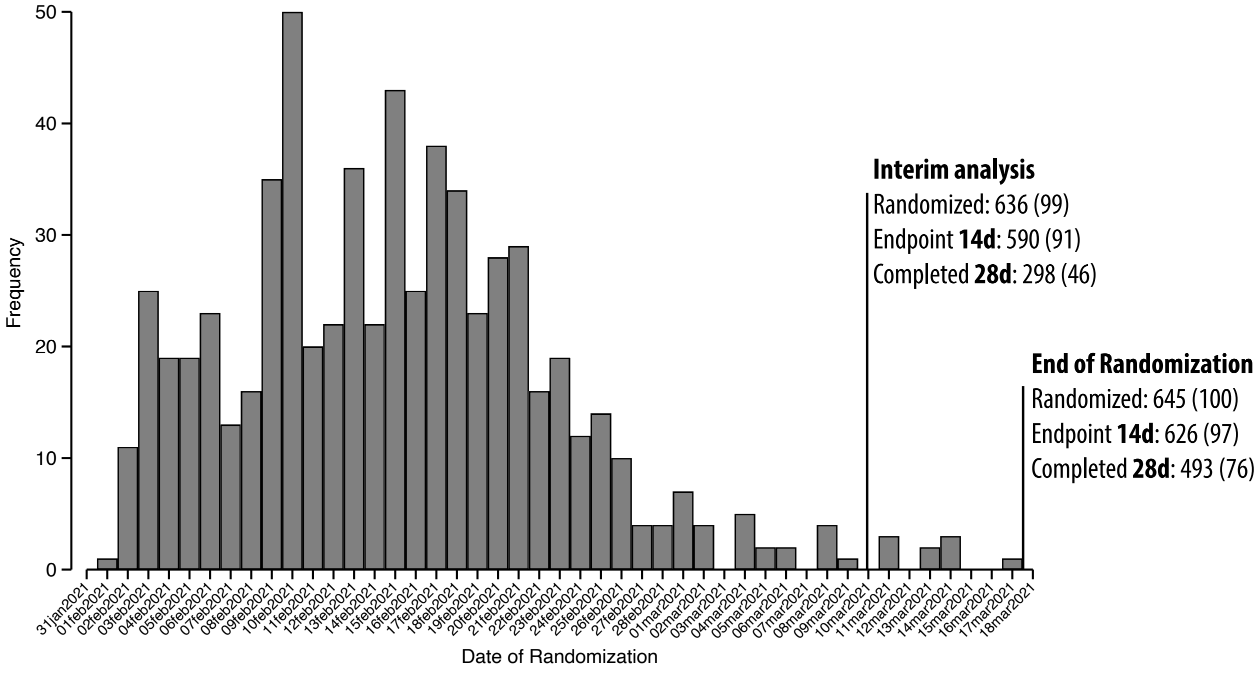

**Figure S1. Randomization/recruitment timeline**

#### Dosage Administration and Compliance Procedures

Proxalutamide 300 mg (3 x 100 mg tablets) or matching placebo was taken orally once daily with or without food, therapy was initiated soon after randomization. Treatment compliance was monitored and recorded while the patient was hospitalized. An accurate and current accounting of the dispensing of the study drug for each subject was maintained on an ongoing basis by the Investigator or delegated personnel. The number of study drug tablets dispensed to the subject was recorded on the Investigational Product Accountability Log. Patients who were discharged before treatment day 14 had the remaining tablets dispensed as to complete the full 14-day treatment course and were actively evaluated for compliance daily until day 14. All centers followed the same protocol.

#### Baseline COVID-19 8-point Ordinal Scale

The protocol was amended before initiation of the trial because of prevailing disease in the participating hospitals. In the original protocol, the inclusion was limited to COVID-19 8-point ordinal scale scores 3, 4, and 5, i.e., did not include patients requiring non-invasive ventilation or high flow oxygen devices.

Prior to the start of the trial, Brazil, and particularly the state of Amazonas, experienced a surge in COVID-19 resulting in an increase in severity of cases. The majority of hospitalized patients on the days preceding the trial were on high flow oxygen devices (COVID-19 8-point ordinal scale score 6). As such, the decision was made to include score 6 on the day of the initiation of recruitment. All patients, including COVID-19 8-point ordinal scale score 6, were randomized in the same 1:1 ratio as indicated in the **Protocol**.

We did not perform randomization stratified per site as a result of batch delivery of drugs or placebo (described above). This led to an unbalance between study arms within hospital sites. In three small hospital centers the randomization strategy had to be modified due to: 1) logistic difficulties in taking the drugs to these remote hospitals and 2) limited financial resources. This was done to avoid unblinding and unmonitored sharing of blister packs among hospitals and patients. We are aware that these changes in the randomization scheme increased the risk of bias in the randomization procedure. However, in the 3 sites located in the large urban city of Manaus with higher hospital certification standards, where randomization could be done as originally planned, a lower mortality rate in both arms was observed, as well as a lower mortality risk ratio (**Table S4**).

**Table S2. Coronavirus disease 2019 8-point ordinal scale scores distribution and outcomes by baseline scores**

| Characteristic | BASELINE SCORES 3-5 | | | BASELINE SCORE 6 | | |
|---|---|---|---|---|---|---|
| **Treatment Group** | **Proxalutamide** N=100 | **Placebo** N=116 | ***Risk ratio (95% CI)*** | **Proxalutamide** N=217 | **Placebo** N=212 | ***Risk ratio (95% CI)*** |
| Females, no. | 52 | 46 | - | 81 | 100 | - |
| Males, no. | 48 | 70 | - | 136 | 112 | - |
| **Recovery over 14 days– n (%)** | **89 (89.0%)** | **62 (53.4%)** | **1.67 (1.39–2.00)** | **169 (77.9%)** | **55 (25.9%)** | **3.00 (2.37–3.80)** |
| Females | 45 (86.5%) | 22 (47.8%) | 1.81 (1.32–2.49) | 64 (79.0%) | 29 (29.0%) | 2.72 (1.97–3.78) |
| Males | 44 (91.7%) | 40 (57.1%) | 1.60 (1.29–2.00) | 105 (77.2%) | 26 (23.2%) | 3.33 (2.35–4.71) |
| **Mortality over 28 days– n (%)** | **5 (5.0%)** | **39 (33.6%)** | **0.15 (0.06–0.36)** | **30 (13.8%)** | **123 (58.0%)** | **0.24 (0.17–0.34)** |
| Females | 1 (1.9%) | 16 (34.8%) | 0.06 (0.01–0.40) | 11 (13.6%) | 55 (55.0%) | 0.25 (0.14–0.44) |
| Males | 4 (8.3%) | 23 (32.9%) | 0.25 (0.09–0.69) | 19 (14.0%) | 68 (60.7%) | 0.23 (0.14–0.36) |
| ***Hazard ratio for death over 28 days (95% CI)*** | 0.13 (0.05-0.33) | | | 0.16 (0.11-0.25) | | |
| **Recovery over 28 days– n (%)** | **93 (93.0%)** | **77 (66.4%)** | **1.40 (1.22–1.61)** | **178 (82.0%)** | **78 (36.8%)** | **2.23 (1.85–2.68)** |
| Females | 50 (96.2%) | 30 (65.2%) | 1.47 (1.19–1.83) | 67 (82.7%) | 41 (41.0%) | 2.02 (1.56–2.60) |
| Males | 43 (89.6%) | 47 (67.1%) | 1.33 (1.10–1.61) | 111 (81.6%) | 37 (33.0%) | 2.47 (1.88–3.25) |
| **Median hospitalization days (IQR)** | 7 (5-10.2) | 11 (8-16) | - | 9 (6-13) | 12 (8-19) | - |
| **Post-randomization time to recovery, Median days (IQR)** | 4 (3-6) | 9 (5-13) | - | 5 (4-8) | 12 (7-17) | - |
| **Day 14 Scores – median (IQR)** | 1 (1-1) | 2 (2-8) | - | 1 (1-2) | 7 (2-8) | - |
| **1.** Not hospitalized, no limitations on activities –no.(%) | 78 (78.0%) | 24 (20.7%) | - | 136 (62.7%) | 8 (3.8%) | - |
| **2.** Not hospitalized, limitation on activities – no. (%) | 11 (11.0%) | 38 (32.8%) | - | 33 (15.2%) | 47 (22.2%) | - |
| **3.** Hospitalized, not requiring supplemental oxygen - no longer requires ongoing medical care – no. (%) | 1 (1.0%) | 5 (4.3%) | - | 2 (0.9%) | 4 (1.9%) | - |
| **4.** Hospitalized, not requiring supplemental oxygen, requiring ongoing medical care – no. (%) | 4 (4.0%) | 5 (4.3%) | - | 7 (3.2%) | 11 (5.2%) | - |
| **5.** Hospitalized, requiring supplemental oxygen – no. (%) | 2 (2.0%) | 4 (3.5%) | - | 9 (4.1%) | 9 (4.2%) | - |
| **6.** Hospitalized, receiving non-invasive ventilation or high flow oxygen devices – no. (%) | 0 (0.0%) | 1 (0.9%) | - | 3 (1.4%) | 5 (2.4%) | - |
| **7.** Hospitalized, on invasive mechanical ventilation – no. (%) | 0 (0.0%) | 7 (6.0%) | - | 4 (1.8%) | 30 (14.2%) | - |
| **8.** Death – no. (%) | 4 (4.0%) | 32 (27.6%) | - | 23 (10.6%) | 98 (46.2%) | - |

#### Table S3. Recovery (coronavirus disease 2019 8-point ordinal scale scores 1 or 2, alive hospital discharge) over 14- and 28-days post-randomization stratified by city

| Characteristic | Proxalutamide | Placebo | *Risk ratio (95% CI)* |
|---|---|---|---|
| **Recovery over 14 days– no. (%)** | 258 (81.4) | 117 (35.7) | 2.28 (1.95-2.66 [P<0.001]) |
| **City** | | | |
| Manaus, Amazonas | 85 (87.6) | 107 (35.9) | 2.44 (2.06–2.89) |
| Manacapuru, Amazonas | 18 (100.0) | - | - |
| Manicore, Amazonas | 5 (100.0) | - | - |
| Maues, Amazonas | 4 (66.7) | 1 (16.7) | 4.00 (0.61–26.1) |
| Itacoatiara, Amazonas | 77 (74.0) | 4 (50.0) | 1.48 (0.73–2.99) |
| Parintins. Amazonas | 69 (79.3) | 5 (31.3) | 2.54 (1.22–5.29) |
| **Recovery over 28 days– no. (%)** | 35 (11.0%) | 162 (49.4%) | 0.22 (0.16-0.31) |
| **City** | | | |
| Manaus, Amazonas | 85 (87.6) | 107 (35.9) | 2.44 (2.06–2.89) |
| Manacapuru, Amazonas | 18 (100.0) | - | - |
| Manicore, Amazonas | 5 (100.0) | - | - |
| Maues, Amazonas | 4 (66.7) | 1 (16.7) | 4.00 (0.61–26.1) |
| Itacoatiara, Amazonas | 77 (74.0) | 4 (50.0) | 1.48 (0.73–2.99) |
| Parintins. Amazonas | 69 (79.3) | 5 (31.3) | 2.54 (1.22–5.29) |

### Table S4. All-cause mortality over 28 days post-randomization stratified by city

| **Characteristic** | **Proxalutamide** | **Placebo** | ***Risk ratio (95% CI)*** |
|---|---|---|---|
| **28-day all-cause mortality– no. (%)** | 35 (11.0) | 162 (49.4) | 0.22 (0.16-0.31 [P<0.001]) |
| **City** | | | |
| Manaus, Amazonas | 6 (6.19) | 145 (48.7) | 0.13 (0.06–0.28) |
| Manacapuru, Amazonas | 0 (0.0) | - | - |
| Manicore, Amazonas | 0 (0.0) | - | - |
| Maues, Amazonas | 2 (33.3) | 5 (83.3) | 0.40 (0.12–1.31) |
| Itacoatiara, Amazonas | 14 (13.5) | 4 (50.0) | 0.27 (0.12–0.63) |
| Parintins. Amazonas | 13 (14.9) | 8 (50.0) | 0.30 (0.15–0.60) |

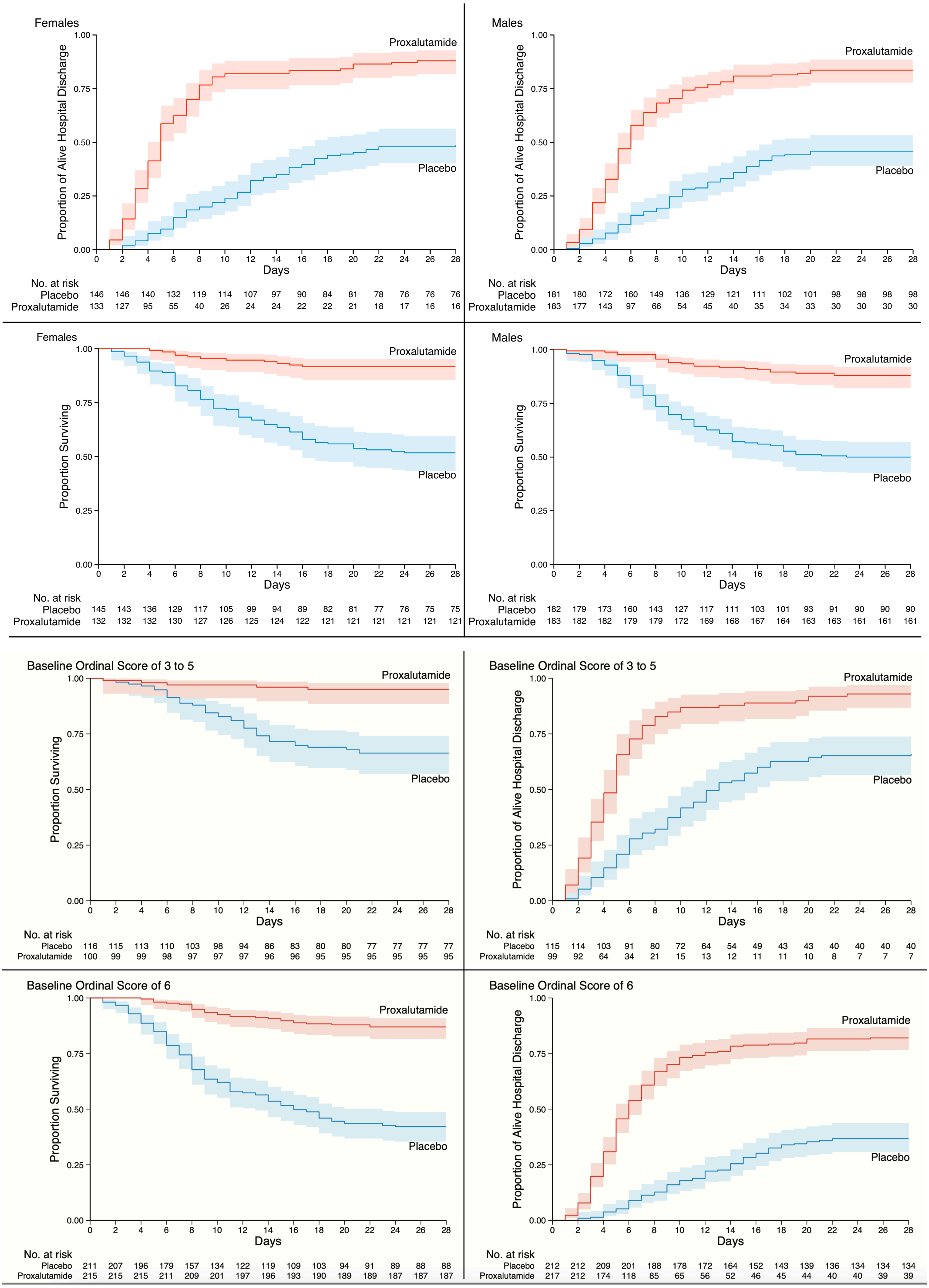

**Figure S2. Kaplan–Meier estimates from randomization to Day 28.**

Alive Hospital Discharge and Proportion Surviving by sex and baseline ordinal scale.

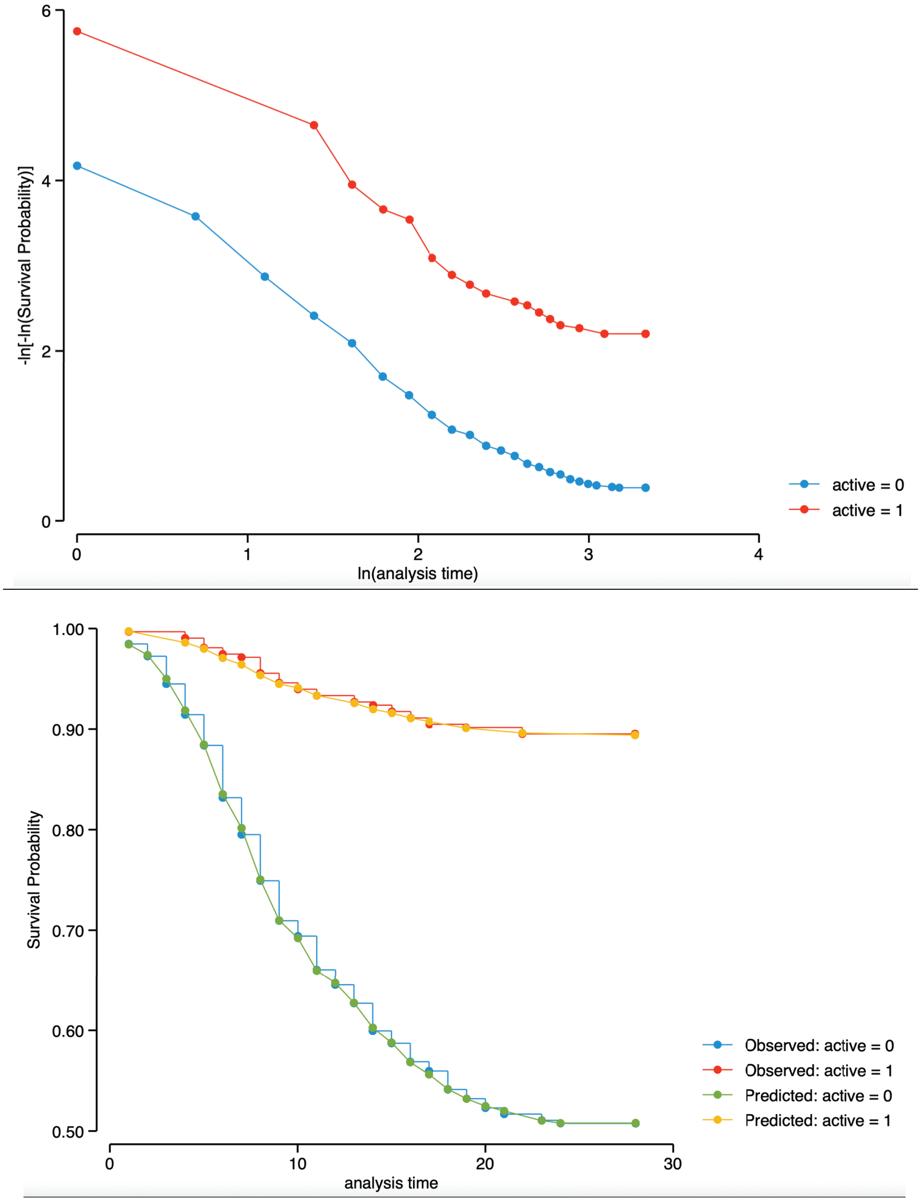

**Figure S3. Graphical assessment of proportional-hazards assumption (hazard ratio over 28 days post-randomization).**

**Top:** figure displays lines that are parallel, implying that the proportional-hazards assumption for the therapy groups has not been violated.

**Bottom:** This is confirmed by Kaplan-Meier versus predicted survival, where the observed values and predicted values are close together for the therapy groups.

Proportional-hazards assumption on the basis of Schoenfeld residuals revealed a global test with P value = 0.7986, therefore, we can assume the proportional hazards.

#### Supplementary Appendix

##### SARS-CoV-2 Lineage Determination

For this post hoc analysis, clinical samples from patients admitted to one of the participating centers testing positive for SARS-CoV-2 in a first RT-qPCR had their samples submitted to a second RT-qPCR performed by BiomeHub (Florianópolis, Santa Catarina, Brazil), using charite-berlin protocol. Only samples with quantification cycle (Cq) below 30 for at least one primer were processed for SARS-CoV-2 genome sequencing by the BiomeHub laboratory.

To perform the SARS-CoV-2 genome sequencing, total RNAs were prepared as in the reference protocol (dx.doi.org/10.17504/protocols.io.befyjbpw) using SuperScript IV (Invitrogen) for cDNA synthesis and Platinum Taq High Fidelity (Invitrogen) for specific viral amplicons. The cDNA obtained were subsequently used for the library preparation with Nextera Flex (Illumina) and quantified with Picogreen and Collibri Library Quantification Kit (Invitrogen). The sequencing was performed on MiSeq 150x150 runs with 500xSARS-CoV-2 coverage (50-100 mil reads/per sample).

The SARS-CoV-2 genome assembly was generated by an in-house pipeline from BiomeHub (Florianópolis, Santa Catarina, Brazil). The remotion of adapters and read trimming in 150 nt were performed with fastqtools.py, followed by the reads mapping to the reference SARS-CoV-2 genome (GenBank accession number NC_045512.2) with Bowtie v2.4.2 (additional parameters: end-to-end and very-sensitive). The mapping coverage and sequencing depth were obtained with samtools v1.11 (minimum base quality per base (Q) $\geq$ 30). Consensus genome sequences were then generated with bcftools mpileup (Q $\geq$ 30; depth (d) $\leq$ 1,000) combined with bcftools filter (DP>50) and bcftools consensus v1.11. Finally, the identification of the SARS-CoV-2 virus lineages was performed by the Pangolin v2.3.8 web server (https://github.com/cov-lineages/pangolin).

#### Table S5. Sequencing of SARS-CoV-2 in 44 Randomly Selected Patients

| Sampling Date | City Center | Lineage (PANGOLIN) |
|---|---|---|
| 18-Feb-2021 | Manaus | P.1 |
| 18-Feb-2021 | Manaus | P.1 |
| 18-Feb-2021 | Manaus | P.1 |
| 18-Feb-2021 | Manaus | P.1 |
| 22-Feb-2021 | Manaus | P.1 |
| 22-Feb-2021 | Manaus | P.1 |
| 22-Feb-2021 | Manaus | P.1 |
| 22-Feb-2021 | Manaus | B.1.1.28 |
| 22-Feb-2021 | Manaus | P.1 |
| 22-Feb-2021 | Manaus | P.1 |
| 22-Feb-2021 | Manaus | P.1 |
| 22-Feb-2021 | Manaus | P.1 |
| 22-Feb-2021 | Manaus | P.1 |
| 22-Feb-2021 | Manaus | P.1 |
| 22-Feb-2021 | Manaus | P.1 |
| 23-Feb-2021 | Manaus | P.1 |
| 23-Feb-2021 | Manaus | P.1 |
| 23-Feb-2021 | Manaus | P.1 |
| 23-Feb-2021 | Manaus | P.1 |
| 23-Feb-2021 | Manaus | P.1 |
| 23-Feb-2021 | Manaus | P.1 |
| 24-Feb-2021 | Manaus | P.1 |
| 24-Feb-2021 | Manaus | P.1 |
| 24-Feb-2021 | Manaus | P.1 |
| 25-Feb-2021 | Manaus | P.1 |
| 25-Feb-2021 | Manaus | P.1 |
| 25-Feb-2021 | Manaus | P.1 |
| 25-Feb-2021 | Manaus | P.1 |
| 25-Feb-2021 | Manaus | P.1 |
| 26-Fev-2021 | Manaus | P.1 |
| 26-Fev-2021 | Manaus | P.1 |
| 01-Mar-2021 | Itacoatiara | P.1 |
| 01-Mar-2021 | Manaus | P.1 |
| 01-Mar-2021 | Manaus | P.1 |
| 01-Mar-2021 | Manaus | P.1 |
| 02-Mar-2021 | Manaus | P.1 |
| 02-Mar-2021 | Manaus | P.1 |
| 03-Mar-2021 | Manaus | P.1 |
| 05-Mar-2021 | Parintins | P.1 |
| 05-Mar-2021 | Manaus | P.1 |
| 08-Mar-2021 | Manaus | P.1 |
| 08-Mar-2021 | Manaus | P.1 |
| 10-Mar-2021 | Manaus | P.1 |
| 10-Mar-2021 | Manaus | P.1 |

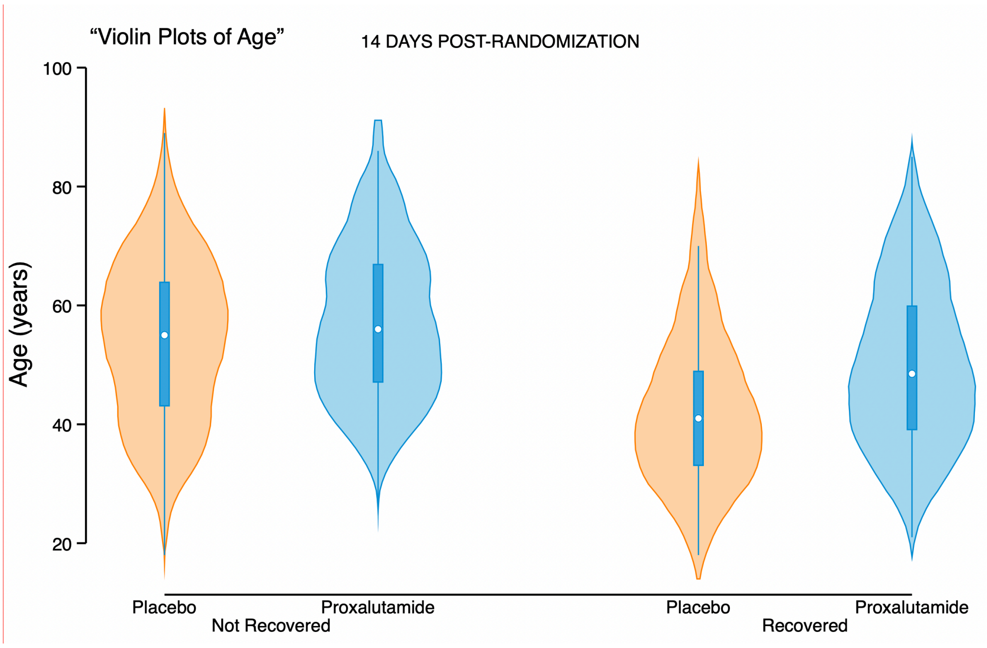

**Figure S4. Violin Plots for Age, for alive hospital discharge over the first 14 days post-randomization, per treatment group.**

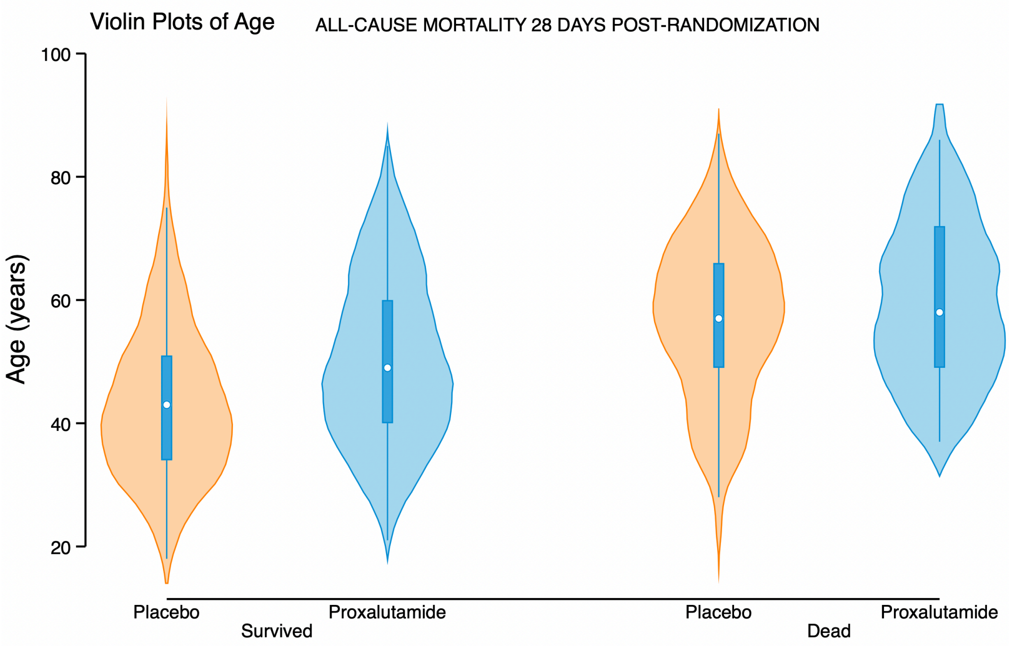

**Figure S5. Violin Plots for Age, for all-cause mortality over the 28 days post-randomization, per treatment group.**

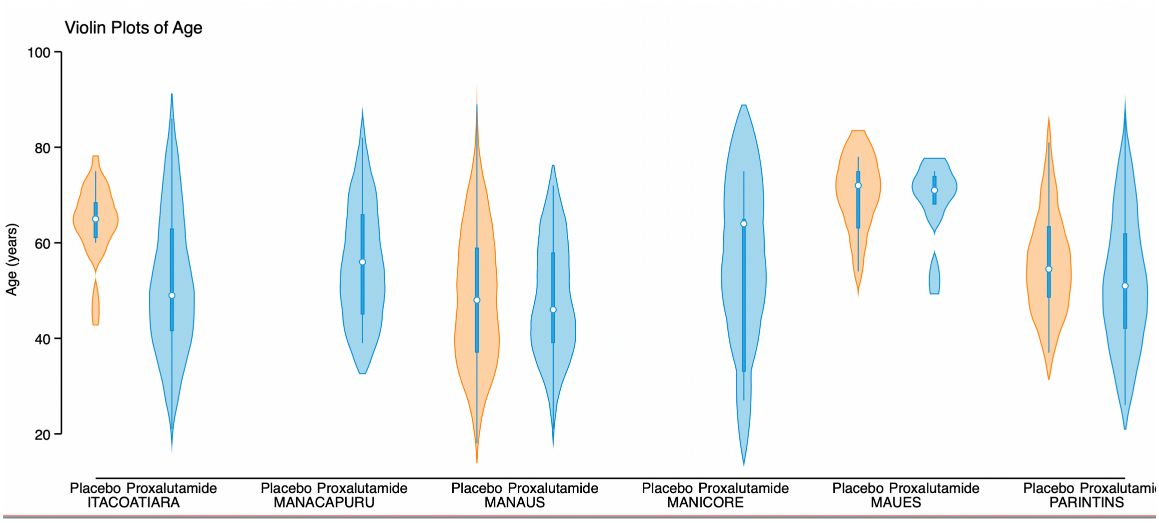

**Figure S6. Violin Plots for Age, for cities, per treatment group.**
